# A multi-task domain-adapted model to predict chemotherapy response from mutations in recurrently altered cancer genes

**DOI:** 10.1101/2023.11.17.23298665

**Authors:** Aishwarya Jayagopal, Robert J. Walsh, Krishna Kumar Hariprasannan, Ragunathan Mariappan, Debabrata Mahapatra, Patrick William Jaynes, Diana Lim, David Shao Peng Tan, Tuan Zea Tan, Jason J. Pitt, Anand D. Jeyasekharan, Vaibhav Rajan

**Author notes:** Corresponding Authors: Dr Anand D. Jeyasekharan. Address: Cancer Science Institute of Singapore, Centre for Translational Medicine (MD6) #13-01G, 14 Medical Drive, Singapore 117599., Dr. Vaibhav Rajan. Address: Department of Information Systems and Analytics, School of Computing, National University of Singapore, Singapore 117417. Both authors contributed equally to the paper (Aishwarya Jayagopal, Robert J. Walsh).

## Abstract

Next generation sequencing (NGS) of tumours is increasingly utilised in oncological practice, however only a minority of patients harbour oncogenic driver mutations benefiting from targeted therapy. Development of a drug response prediction (DRP) model based on available genomic data is important for the ‘untargetable’ majority of cases. Prior DRP models typically rely on whole transcriptome and whole exome sequencing (WES), which is often unavailable in clinical practice. We therefore aim to develop a DRP model towards repurposing of standard chemotherapy, requiring only information available in clinical grade NGS (cNGS) panels of recurrently mutated genes in cancer. Such an approach is challenging due to the sparsity of data in a restricted gene set and limited availability of patient samples with documented drug response. We first show that an existing DRP performs equally well with whole exome data and a cNGS subset comprising ∼300 genes. We then develop Drug IDentifier (DruID), a DRP model specific for restricted gene sets, using a novel transfer learning-based approach combining variant annotations, domain-invariant representation learning and multi-task learning. Evaluation of DruID on pan-cancer data (TCGA) showed significant improvements over state-of-the-art response prediction methods. Validation on two real world - colorectal and ovarian cancer - clinical datasets showed robust response classification performance, suggesting DruID to be a significant step towards a clinically applicable DRP tool.

## Introduction

Precision oncology has shifted treatment paradigms in solid organ tumours over recent years, underpinned by widespread adoption of somatic next generation sequencing (NGS) and increasing knowledge of molecular aberrations present within tumours. However, only a minority of patients undergoing NGS go on to receive biomarker directed, or ‘matched’, treatment, which currently follows a single gene, single target approach with oncogenic drivers such as *EGFR, NTRK, RET* and *BRAF* (Tsimberidou 2019). There remains an unmet need to better tailor or repurpose treatment for the majority of patients who lack such genomic targets based on clinical grade NGS (cNGS). Drug Response Prediction (DRP) models utilising machine learning (ML) to predict therapeutic responses represent an appealing solution.

Numerous deep learning strategies have been published in recent years using available multi-omics data from cell line, patient-derived xenograft (PDX) and patient datasets (He 2022, Jia 2021, Partin 2023). Cancer cell lines provide the majority of ground truth drug response data for such endeavours (Adam 2020, Chen 2021, Firoozbakht 2021) however, DRP models trained on cell lines alone often translate poorly to patients (Mourragui 2019, Mourragui 2021, Sharifi-Noghabi 2020). This is partly due to inherent biological differences, meaning cell lines do not accurately represent patient tumours. Cell lines are essentially a subpopulation of the primary tumour and do not exhibit heterogeneity seen *in vivo*. The absence of the tumour microenvironment and interactions with the host of stromal cells present in patients is also key (Mourragui 2019, Huo 2020). In addition, technical differences in response measurement in cell lines versus in patients, and differences in drug dosing between cell lines and patients will affect interpretation of results by a DRP model.

While omics data is increasingly available for many cancer patients (TCGA 2013, Cerami 2012), drug response data for these patients remains scarce and limited to standard of care therapies only. To address such challenges, transfer learning approaches including domain adaptation have been developed to train DRP models from both cell lines and patients (He 2022, Sharifi 2021, Ma 2021).

Prior studies have used omics data from 4 categories - genomics (mutation, copy number variation (CNV)), transcriptomics (gene expression microarrays, RNA-seq), epigenomics (methylation) and proteomics (Reverse Phase Protein Arrays (RPPA)) (Partin 2023). While studies on cell lines have shown gene expression data to outperform mutations (Costello 2014, Levatic 2022), recent studies on patients have also identified the relevance of mutations in determining survival outcomes (Liu 2022). State-of-the-art transfer learning methods, which evaluated their models on patient data, have largely restricted their analysis to gene expression data (Sharifi 2021, Peres 2021). The genes selected in these methods are not captured based on their presence in cNGS panels; nor are the number of chosen genes comparable across cNGS and these methods. For example, CODE-AE(He 2022) used a set of 1426 genes, which showed the most variation in gene expression values, and Velodrome(Sharifi 2021) used a set of 2128 genes, which were chosen based on known molecular interactions amongst proteins. Even when these transfer learning methods (He 2022) used mutations or combinations of mutations and gene expression, they reported better performance with gene expression. Requiring transcriptomic input data represents a challenge in bringing these methods to mainstream patient care and it remains unknown if such tools can accurately predict response from the limited number of recurrently altered cancer genes that are included in cNGS panels such as FoundationOne CDx (324 genes), Tempus (523 genes), and TruSight Oncology 500 (523 genes). To the best of our knowledge, no prior transfer learning methods have been evaluated on such a restricted subset of genes. Moreover, methods which have used mutations as inputs, have not considered the variant level information captured in cNGS reports; instead they treat all alterations as equal, resulting in loss of granularity and potential reduction in predictive accuracy (Table 2).

In this paper, we make two contributions - (1) we evaluate the efficacy of extant DRP methods on the limited subset of genes available in cNGS panels and (2) we develop a new model, called Drug IDentifier (DruID), specifically designed for use with cNGS panels and address the modelling challenges posed by such data. We first compare the performance of DRP models CODE-AE(He 2022) and Velodrome (Sharifi 2021) on subsets of genes from cNGS panels against an extended gene list from whole exome sequencing data (WES with 19,536 genes). Although cNGS panels show no significant difference in performance compared to WES, the DRP performance itself is low for all panels. We attribute the inferior performance of existing methods to their inadequate modelling of sparse mutation data and neglecting the fine-grained variant level information available in cNGS reports. We addressed these limitations by designing DruID. DruID leverages advanced deep learning and transfer learning techniques and a novel multi-stage approach comprising variant annotation-based feature engineering, unsupervised generative modelling and supervised multi-task learning. DruID utilised both (a) fine-grained variant information in relatively abundant unlabelled (without drug response information) cell line and patient data, and (b) limited labelled (with drug responses) patient data. The training procedure of DruID is carefully designed to account for both differences in mutation distribution and drug response across the domains of cell lines and patients.

DruID is shown to outperform existing state of the art DRP models in predicting response in a cohort of patients from The Cancer Genome Atlas (TCGA). Using clinical datasets from a tertiary oncology centre in Singapore, DruID’s performance is validated in patients with advanced colorectal and ovarian cancer, with robust response prediction seen on these clinical cohorts.

## Results

### Datasets used in this study

Four datasets were used in this study: DepMap (v2021Q3), TCGA, and two cancer-specific datasets, IMAC-OV and IMAC-CRC, containing patients with advanced ovarian and colorectal cancer respectively (Table 1). The cancer-specific datasets (IMAC-OV and IMAC-CRC) were collected as part of the ongoing Integrated molecular analysis of cancer (IMAC) and IMAC-Gynaecologic Oncology (IMAC-GO) studies from the National University Cancer Institute, Singapore (NCIS). The detailed inclusion and exclusion criteria, data pre-processing and experimental procedures are documented in Methods. The mutational information present in IMAC-OV and IMAC-CRC cohorts was obtained using the FoundationOne CDx panel (324 genes); we conducted the majority of our experiments using only the genes available in this panel. We evaluated all DRP models on a subset of drugs with a sufficiently large number of recorded responses in patients.

**Table 1:** Overview of datasets used.

|  | DepMap | TCGA | IMAC-OV | IMAC-CRC |
| --- | --- | --- | --- | --- |
| ***N*** | 689 | 470 | 105 | 82 |
| **Age, median (min-max)** | NA | 59 (24-85) | 59.7 (25-81) | 59 (37-83) |
| **Gender Female (Male)** | NA | 54.9%<br>(45.1%) | 100% | 41.5%<br>(47.3%)<br>[missing info<br>- 1 patient] |
| **Primary Site** |  |  |  |  |
| Ovary/fallopian tube/peritoneum | 34 (4.9%) | 0 | 105 (100%) | 0 |
| Colon & Rectal | 43 (6.2%) | 34(7.2%) | 0 | 82 (100%) |
| Others (lung, stomach, head & neck, bladder, skin, uterus, breast, cervical, brain, pancreas, oesophageal, liver, prostate etc) | 612 (88.8%) | 436 (92.8%) | 0 | 0 |
| **Treatment** |  |  |  |  |
| Cisplatin/Carboplatin | 537 (77.9%) | 206 (43.8%) | 105 (100%) | 0 |
| Paclitaxel | 676 (98.1%) | 113 (24%) | 102 (97.1%) | 0 |
| 5-Fluorouracil | 589 (85.5%) | 125 (26.6%) | 0 | 82 (100%) |
| Irinotecan | 668 (97%) | 0 | 0 | 30 (36.6%) |
| Oxaliplatin | 555 (80.6%) | 0 | 0 | 51 (62.2%) |
| **Omics availability** |  |  |  |  |
| Mutation | Yes | Yes | Yes | Yes |
| Copy Number | Yes | Yes | Yes | Yes |
| Gene Expression | Yes | Yes | No | No |

Details of the datasets used in this paper, including source from where each dataset was obtained, the type of cancer in each dataset, the number of available patients(samples for cell lines) and set of drugs administered to these patients, obtained after data preprocessing. *Abbrev.* TCGA, The Cancer Genome Atlas; CCLE, cancer cell line encyclopaedia; IMAC, Integrated molecular analysis of cancer; OV, ovarian cancer; CRC, colorectal cancer

**Table 2:**
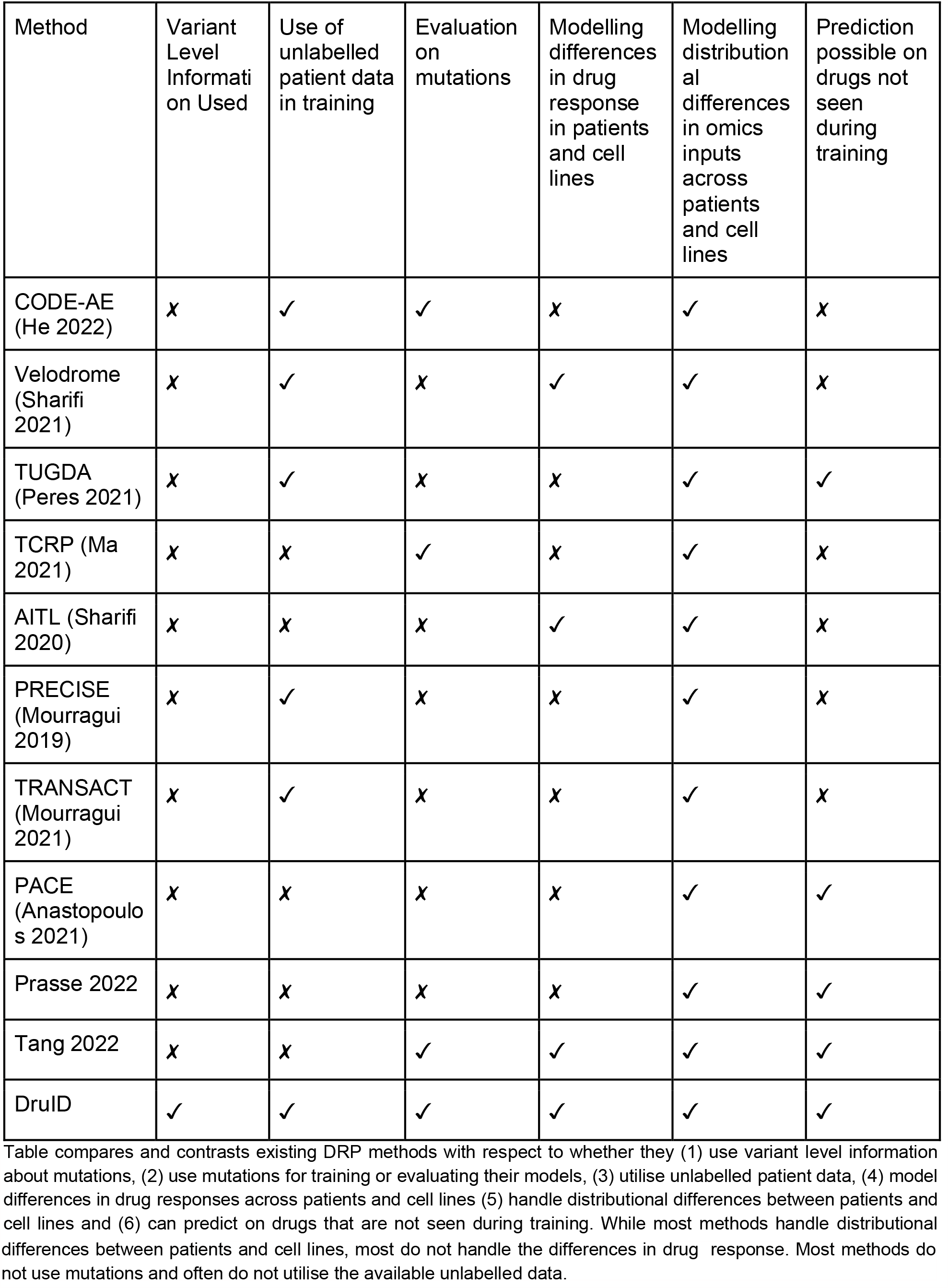
Comparison of existing DRP methods and DruID.

| Method | Variant Level Information Used | Use of unlabelled patient data in training | Evaluation on mutations | Modelling differences in drug response in patients and cell lines | Modelling distributional differences in omics inputs across patients and cell lines | Prediction possible on drugs not seen during training |
| --- | --- | --- | --- | --- | --- | --- |
| CODE-AE (He 2022) | x | ✓ | ✓ | x | ✓ | x |
| Velodrome (Sharifi 2021) | x | ✓ | x | ✓ | ✓ | x |
| TUGDA (Peres 2021) | x | ✓ | x | x | ✓ | ✓ |
| TCRP (Ma 2021) | x | x | ✓ | x | ✓ | x |
| AITL (Sharifi 2020) | x | x | x | ✓ | ✓ | x |
| PRECISE (Mourragui 2019) | x | ✓ | x | x | ✓ | x |
| TRANSACT (Mourragui 2021) | x | ✓ | x | x | ✓ | x |
| PACE (Anastopoulos 2021) | x | x | x | x | ✓ | ✓ |
| Prasse 2022 | x | x | x | x | ✓ | ✓ |
| Tang 2022 | x | x | ✓ | ✓ | ✓ | ✓ |
| DrulD | ✓ | ✓ | ✓ | ✓ | ✓ | ✓ |

Table compares and contrasts existing DRP methods with respect to whether they (1) use variant level information about mutations, (2) use mutations for training or evaluating their models, (3) utilise unlabelled patient data, (4) model differences in drug responses across patients and cell lines (5) handle distributional differences between patients and cell lines and (6) can predict on drugs that are not seen during training. While most methods handle distributional
differences between patients and cell lines, most do not handle the differences in drug response. Most methods do not use mutations and often do not utilise the available unlabelled data.

### DRP models based on clinical NGS data perform sufficiently accurate

We first wished to evaluate if drug response prediction models could be generated from mutational information on a subset of genes that are recurrently mutated in cancer, and captured by clinical grade NGS panels. To do this, we compared the performance of CODE-AE (He 2022), a state-of-the-art transfer learning approach, on mutation data from three subsets of genes. Subset1 comprises 324 genes included in FoundationOne CDx analyses (Foundation Medicine, Cambridge MA), while Subset2 consists of 285 genes common across FoundationOne CDx, TruSight Oncology 500(Illumina, San Diego, CA) and Tempus xF+ (Tempus XF+) cNGS panels. Subset3 includes 19,536 genes, nearly all those available from WES.

Pan-cancer data from TCGA (Table 1) was used to evaluate CODE-AE performance on 3 drugs (5-Fluorouracil, Cisplatin and Paclitaxel) where sufficient samples (patient, drug pairs) were available. Three train-test splits were created by random sampling. In each split, CODE-AE was trained on cell line data from the cancer cell line encyclopaedia (CCLE) and TCGA training set, and evaluated on the corresponding TCGA test set. We evaluated performance in classifying responders (categories complete [CR] or partial response [PR] by Response Evaluation Criteria In Solid Tumours (RECIST) v1.1 criteria) from non-responders (stable [SD] or progressive disease [PD]). In total the test set had 203 samples (patient-drug pairs), with 90, 82 and 90 pairs across the three splits.

The classification performance of CODE-AE for the three gene subsets is shown in Fig. 1. Area under receiver operating characteristics curve (AUROC) and area under precision recall curve (AUPRC) are comparable (Fig. 1.a), with no significant difference between gene subsets (*p* > 0.05, ANOVA). Figure 1.b shows the confusion matrices at a specific, arbitrarily chosen threshold (false positive rate = 0.3, true positive rate = 0.3). Subset3 (WES gene panel) enabled identification of more responders than the Subset1 and Subset2 (47 vs 41 and 47 vs 44, respectively). Specificity, precision and sensitivity metrics are equivalent across gene subsets (Supplementary Table 1). Subset3 has the highest accuracy (specificity = 0.692, sensitivity = 0.311, precision = 0.746), followed by Subset2 (specificity = 0.692, sensitivity = 0.291, precision = 0.733) and Subset1 (specificity = 0.673, sensitivity = 0.272, precision = 0.707). A similar comparison, using another DRP, Velodrome, also showed no significant difference in AUROC and AUPRC between input gene subsets (supplementary Fig 2), suggesting that information from limited gene panels is sufficient to build a DRP model of similar accuracy to that from WES gene panels.

**Figure 1.**
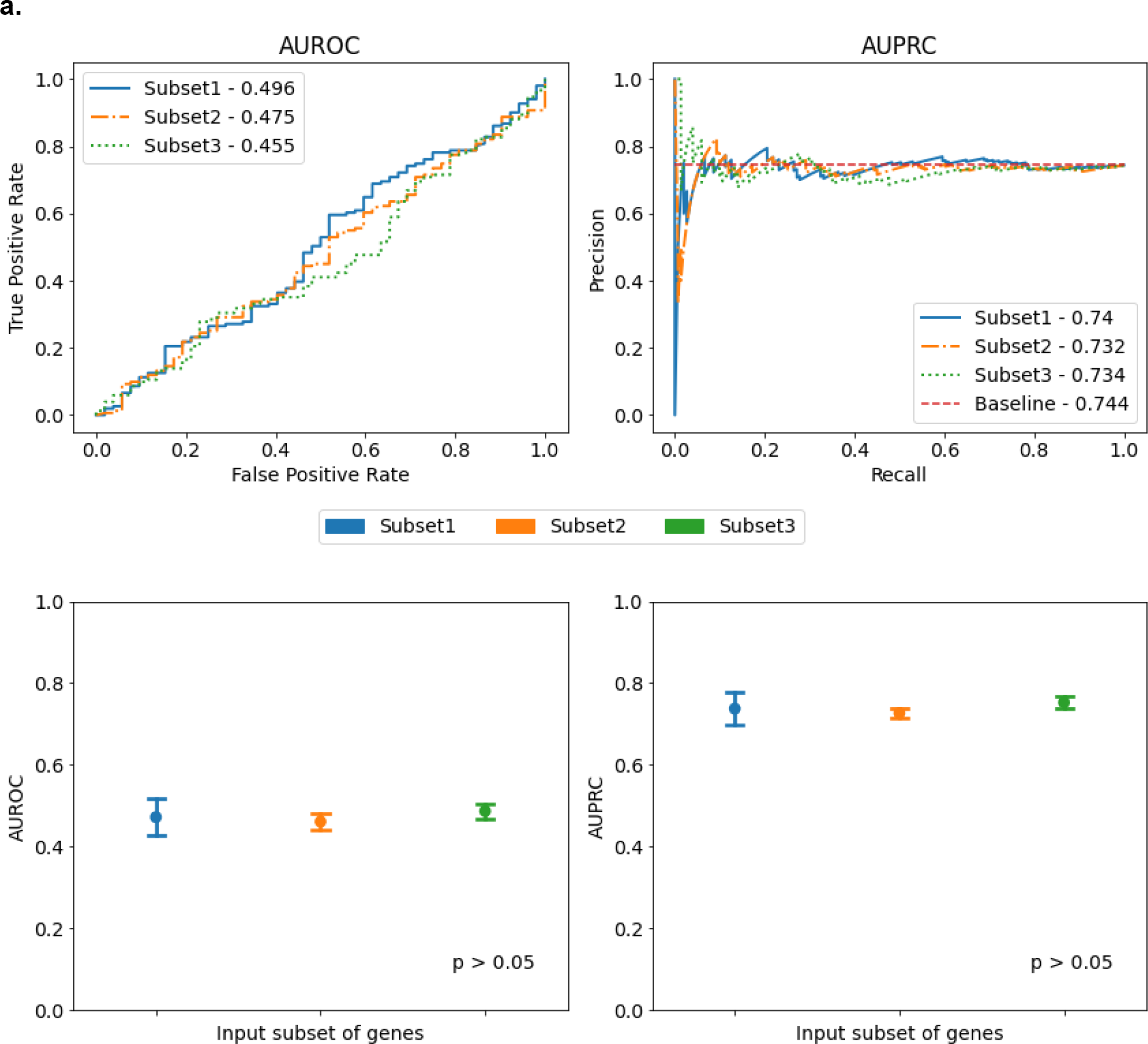
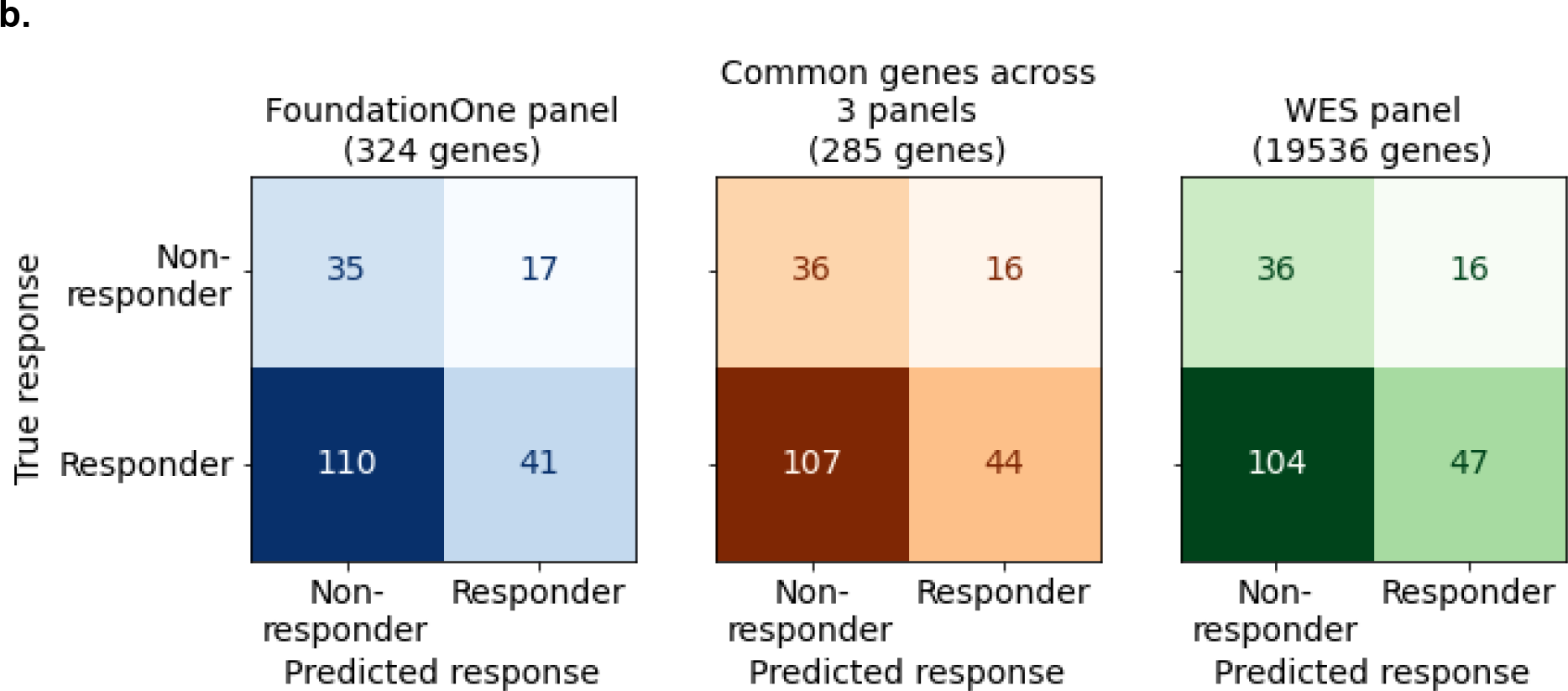
**(a):** Comparison of AUROC (area under receiver operating characteristics; top left panel) and AUPRC (area under precision recall curve; top-right panel) scores of response prediction using CODE-AE with different input subsets of genes. Baseline AUPRC is the fraction of positive labelled test (patient, drug) pairs with respect to all test (patient, drug) pairs. Performance(bottom panels) is measured over 3 randomly chosen test splits (mean ± S.E.M). Significance is assessed by Anova. (**b)**. Confusion matrices for different input subsets of genes on 203 samples (patient-drug pairs) from TCGA, predictions obtained using the method CODE-AE. Colour indicates the input subset, shade indicates magnitude of the values.

### DruID: an improved model for predicting chemotherapy drug response with cNGS data

As seen in Fig. 1, cNGS panels with limited subsets of genes have a predictive power similar to that of a WES panel. However, it can also be observed that when using CODE-AE and Velodrome (Supplementary Fig. 2), the overall performance is quite poor (AUROC < 0.5 and AUPRC < baseline). This alludes to the need for building better predictive DRP models. We attributed this performance to (1) inadequate modelling of sparse mutation data and (2) loss of granularity by not utilising variant information available in cNGS panels. In this section, we introduce DruID - a novel transfer learning-based drug identifier model which addresses both these issues.

#### DruID: Model Overview

There are two challenges, in building DRP models using data from cNGS panels, that we address. The first challenge arises due to sparsity in the input data. Most patients have just a few mutations among the panel of genes considered, which leads to highly sparse input features. For example, consider the FoundationOne CDx panel with 324 genes and a simple one-hot vector feature representation indicating presence/absence of mutations. In such a case, each patient would be represented by a 324-dimensional binary vector which would typically have very few non-zero values. Moreover, if additional features are used per gene, the number of coordinates per gene increases and sparsity may increase further.

The second challenge arises due to limited labelled patient data for training. Previous works have utilised preclinical data (drug responses on cell lines) through domain adaptation techniques to address this challenge(He 2022, Sharifi 2021, Peres 2021). Cell lines and patients are considered as two distinct domains as both the distributions of mutations and response to drugs differ across these two domains. The measurements of responses also differ – real-valued Area Under Dose Response Curve (AUDRC) or Half Maximal Inhibitory Concentration (IC50) values for cell lines and categorical Response Evaluation Criteria In Solid Tumours (RECIST) scores for patients. While labelled patient data is limited, the number of unlabelled patient data samples (i.e., without drug responses) is much higher, and can be utilised during training, for a suitably designed model.

Our model, DruID, addresses these challenges through a novel synthesis of machine learning techniques. Figure 2(a) shows the three stages of DruID:

i. Variant annotations
ii. Unsupervised domain-invariant representation learning
iii. Multi-task Drug Response Prediction

**Figure. 2.**
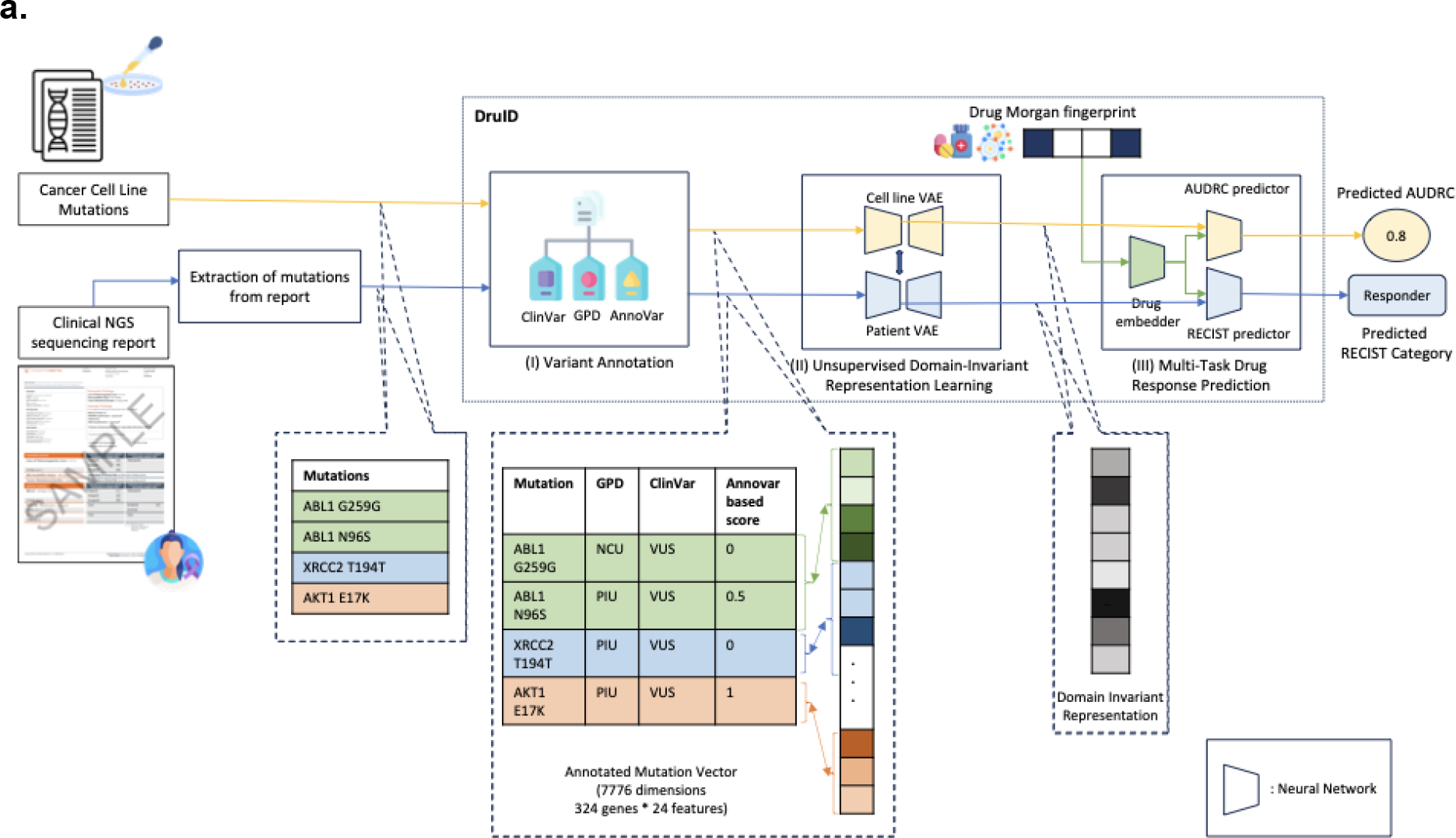
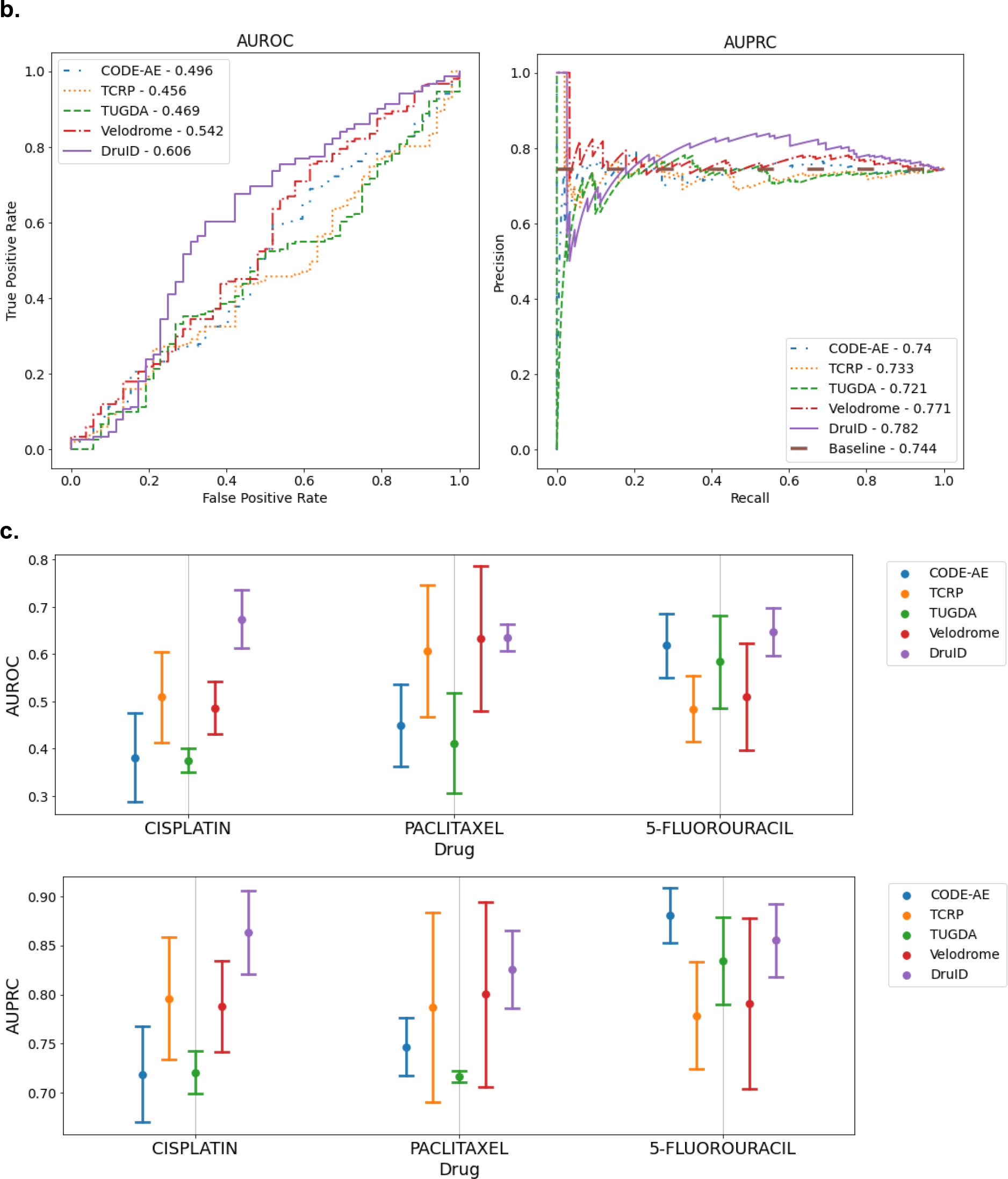
(a): Overview of DruID. During training, DruID takes as input the set of mutations available from cell lines as well as from the clinical NGS(cNGS) sequencing reports of patients (pathogenic variants and variants of uncertain significance). The mutations from both cell lines and patients are passed through Stage (I) Variant Annotation. ClinVar, Gene-to-Protein-to-Disease (GPD) and Annovar are used to obtain annotations for each mutation. GPD returns one of 3 categories - protein information unit(PIU), linker unit(LU) or non-coding unit(NCU) for each mutation. ClinVar returns one of 3 categories - benign, pathogenic or variant of unknown significance(VUS). Annovar returns predictions indicating deleteriousness of a variant, from 17 algorithms, which are averaged to obtain a score. Next, all mutations in the same gene are aggregated over all 3 GPD categories and 3 ClinVar categories, using mean, max, sum and count operations. These are further concatenated to obtain a 7776-dimensional annotated mutation vector. In Stage (II) Unsupervised Domain-Invariant Representation Learning, the annotated mutation vectors of cell lines and patients are passed through two separate VAEs to obtain lower dimensional representations. An additional alignment is done to ensure domain-invariant representations are learnt. The VAEs also use Zero-inflated distributions to model sparse data. The learnt representations along with drug Morgan fingerprints are passed to Stage (III) Multi-task drug response prediction which predicts AUDRC score for cell lines and RECIST category for patients. (**b):** Performance of DruID and comparator methods on response prediction from TCGA patient cohort. Left: AUROC of 5 drug response prediction (DRP) methods. Right: AUPRC of 5 DRPs. Baseline AUPRC - 0.744. (**c):** Comparison of response prediction for each drug. Mean AUROC (above), and mean AUPRC (below) across 3 test splits with standard error corresponding to each drug.

In stage I, we design features (or numeric representations of the inputs) based on various functional annotations that provide fine-grained variant-specific information (Landrum 2017, Li 2020, Wang 2010). This enables us to fully utilise the information available in cNGS panels. To the best of our knowledge, no previous approach has used variant-level information for drug response prediction.

In stage II, numeric representations of cell lines and patients are used together to obtain another low-dimensional domain-invariant representation. This stage has multiple goals. Since mutation-based representations are extremely sparse, we use Zero-inflated distributions to model them. Further, we use Variational Autoencoders (VAEs) which are specialised neural models, based on generative artificial intelligence (AI), to obtain dense lower-dimensional representations. Separate VAEs for cell lines and patients are used to model distinct distributions. They are trained together to align their lower-dimensional representations such that their distributional characteristics are similar across the domains – these domain-invariant representations are then used to train a multi-task drug response prediction model in stage III.

In stage III, a neural model is trained to predict both AUDRC for cell lines (a regression task) and response categories (responders [PR or CR] or non-responders [SD or PD]) in patients (a binary classification task) for a given input drug. The Morgan fingerprint of the drug is used as an additional input. The model is designed to simultaneously train on these two tasks which enables both sharing of information across the two tasks and task-specific modelling in cell lines and patients, accounting for differences in their drug responses.

To validate the importance of each of the components in DruID, we conducted an ablation study by removing the variant annotation stage first, followed by the modification of the VAEs to exclude the Zero-inflated distribution (Supplementary Fig. 3). We observe that there is a reduction in both AUROC and AUPRC with the removal of each component, indicating their importance in the overall performance.

Our modelling approach has several advantages. Since stage II is unsupervised we can utilise large amounts of available unlabelled patient data to obtain accurate representations of patient data. Stages II and III can be first trained on pan-cancer data and then fine tuned on input specific to a cancer type and/or drug to obtain cancer and/or drug specific models. By using drug fingerprints as inputs in stage III, the model can predict on drugs not seen during training - important for potential applications in drug repurposing or discovery. Finally, the VAE in stage II can be extended to model multimodal data (Mariappan 2022) including additional genomic or transcriptomic inputs. Refer to Methods for further details comparing DruID against other DRP methods.

#### DruID improves response prediction results

We evaluate the performance of DruID and four other transfer learning-based approaches – CODE-AE (He 2022), Velodrome (Sharifi 2021), TCRP (Ma 2021) and TUGDA (Peres 2021) – on TCGA.

Figure 2(b) shows the ROC and PRC curves along with the AUROC and AUPRC values of all the methods. DruID achieves the highest AUROC and AUPRC values of 0.606 and 0.782 respectively, while Velodrome is the only other DRP to achieve AUROC and AUPRC values above the respective baselines of 0.5 and 0.744. The performance of DruID is significantly better than that of Velodrome, in terms of both AUROC (*p*=0.004) and AUPRC (*p*=0.037).

Performance for each of three drugs, cisplatin, paclitaxel and 5-fluorouracil (5-FU) is shown in Fig. 2(c). DruID performs consistently across the compounds and is the only model to achieve AUROC above the 0.5 baseline for each drug, while other methods show more variations in performance. For Cisplatin, DruID has the highest average AUROC (*p*=0.111 compared to TCRP). For Paclitaxel, AUROC of DruID and Velodrome are comparable(p=0.496) but the variance of Velodrome is much higher. For 5-FU, performance of DruID and CODE-AE are comparable(*p*=0.371). With respect to precision-recall, for Cisplatin, DruID has the best mean AUPRC followed by that of TCRP(*p*=0.212). For Paclitaxel, DruID has the highest mean AUPRC(*p*=0.408 compared to Velodrome). For 5-FU, CODE-AE has the highest mean AUPRC, followed by DruID. The difference in p-value was not significant in these cases.

### Inclusion of copy number variation (CNV) or gene expression data does not improve DruID performance

We next evaluated the effect of including copy number variations (CNV) on model performance, which is available in many cNGS panels. The data was used directly, unlike mutation data where features based on variant annotations were used. For CNV of each gene, we had a count value indicating amplification, loss or no change. DruID allows us to model such data using Zero-inflated Negative Binomial distributions within the VAE training (detailed in Methods).

TCGA was used for evaluation. We compared the performance of DruID on 3 different input types: annotated mutations, combined CNV and annotated mutations, and CNV alone. In all 3 cases, only 324 genes represented in FoundationOne CDx were used and performance was measured over 3 test splits. Results are seen in Fig. 3(a) with DruID’s predictive performance with annotated mutations alone shown to be significantly better than annotated mutations with CNV information, both in terms of mean AUROC (*p* = 0.003) and AUPRC (*p* = 0.013) over the 3 test splits.

**Figure. 3.**
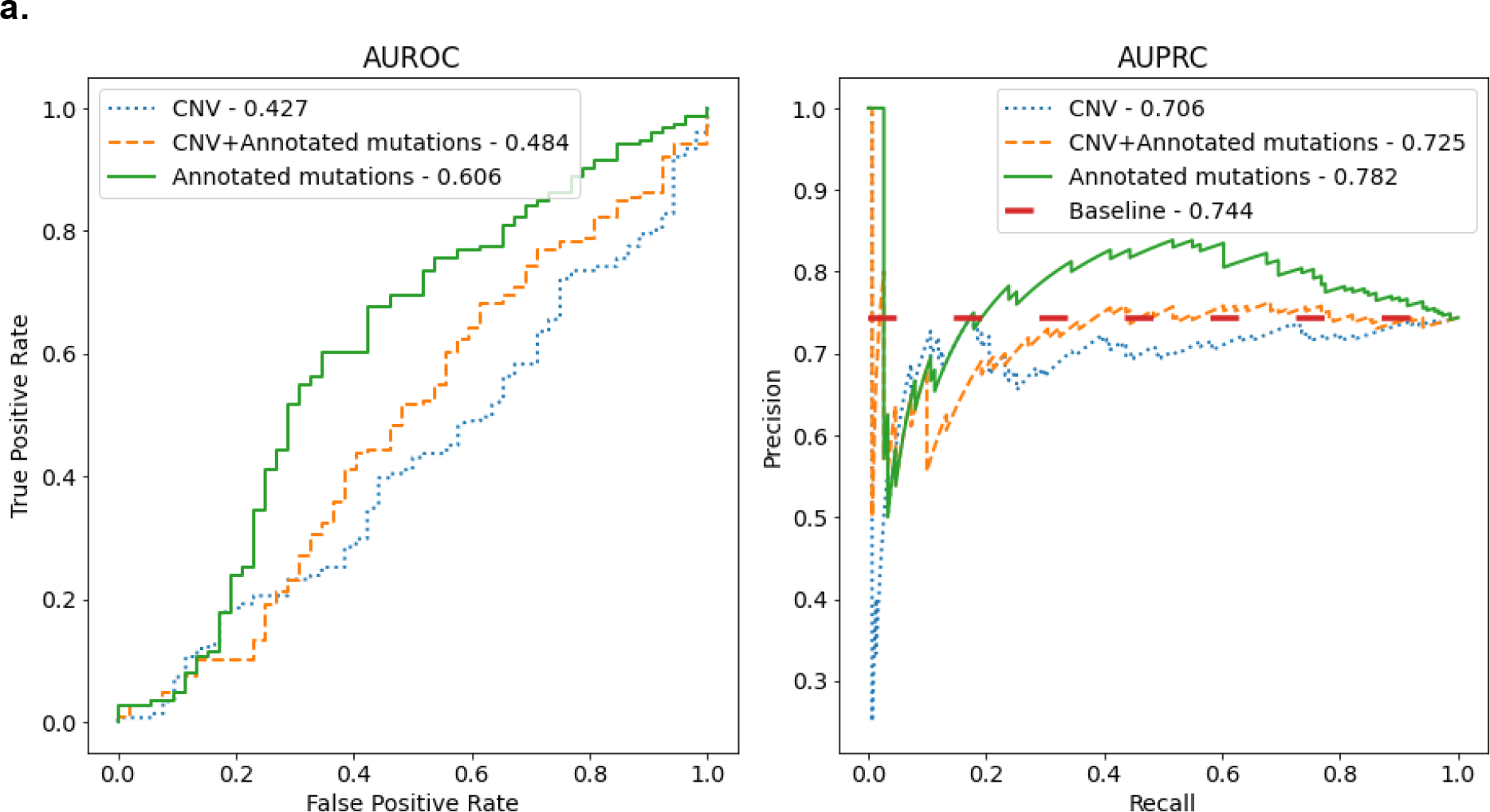
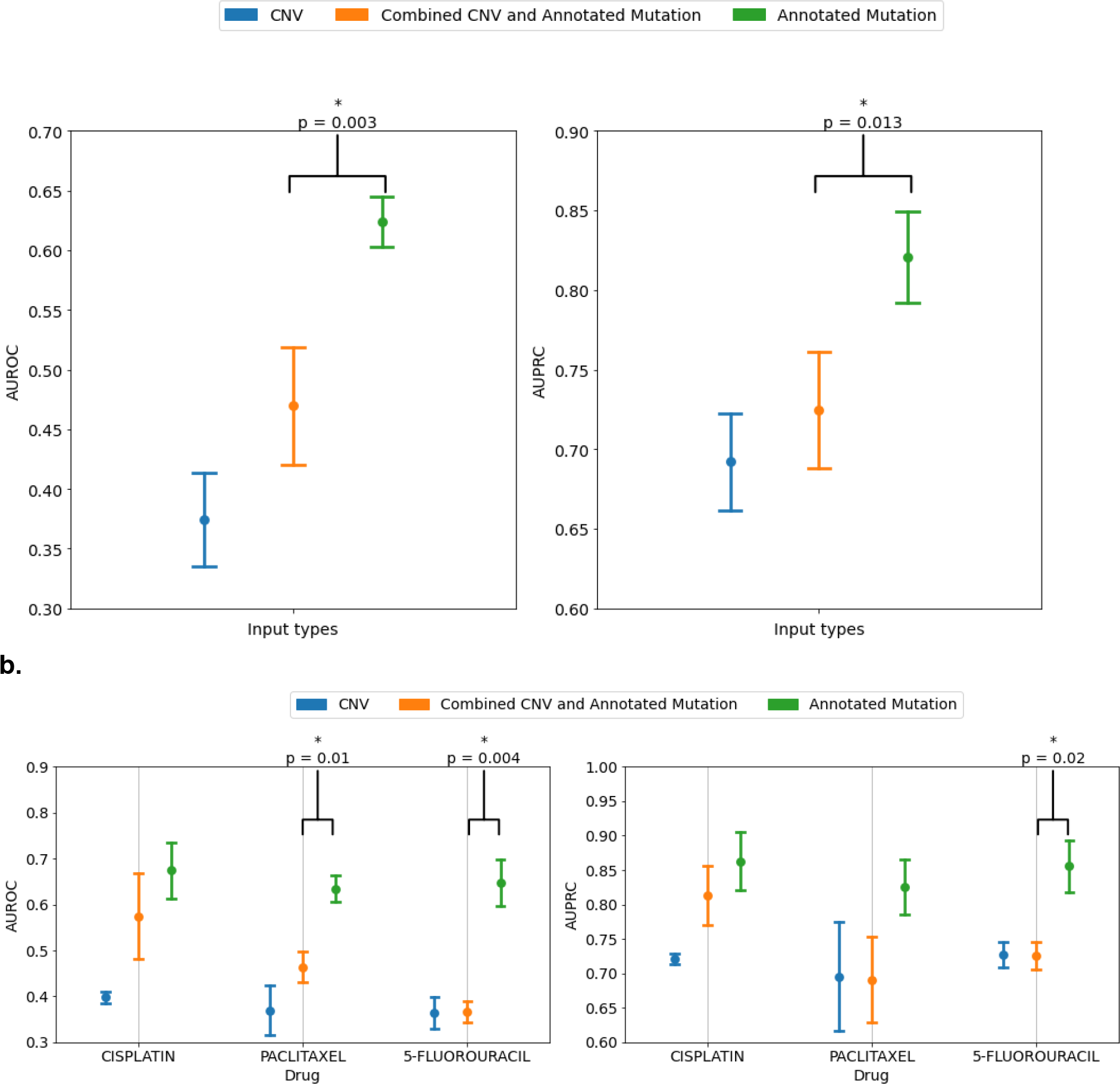
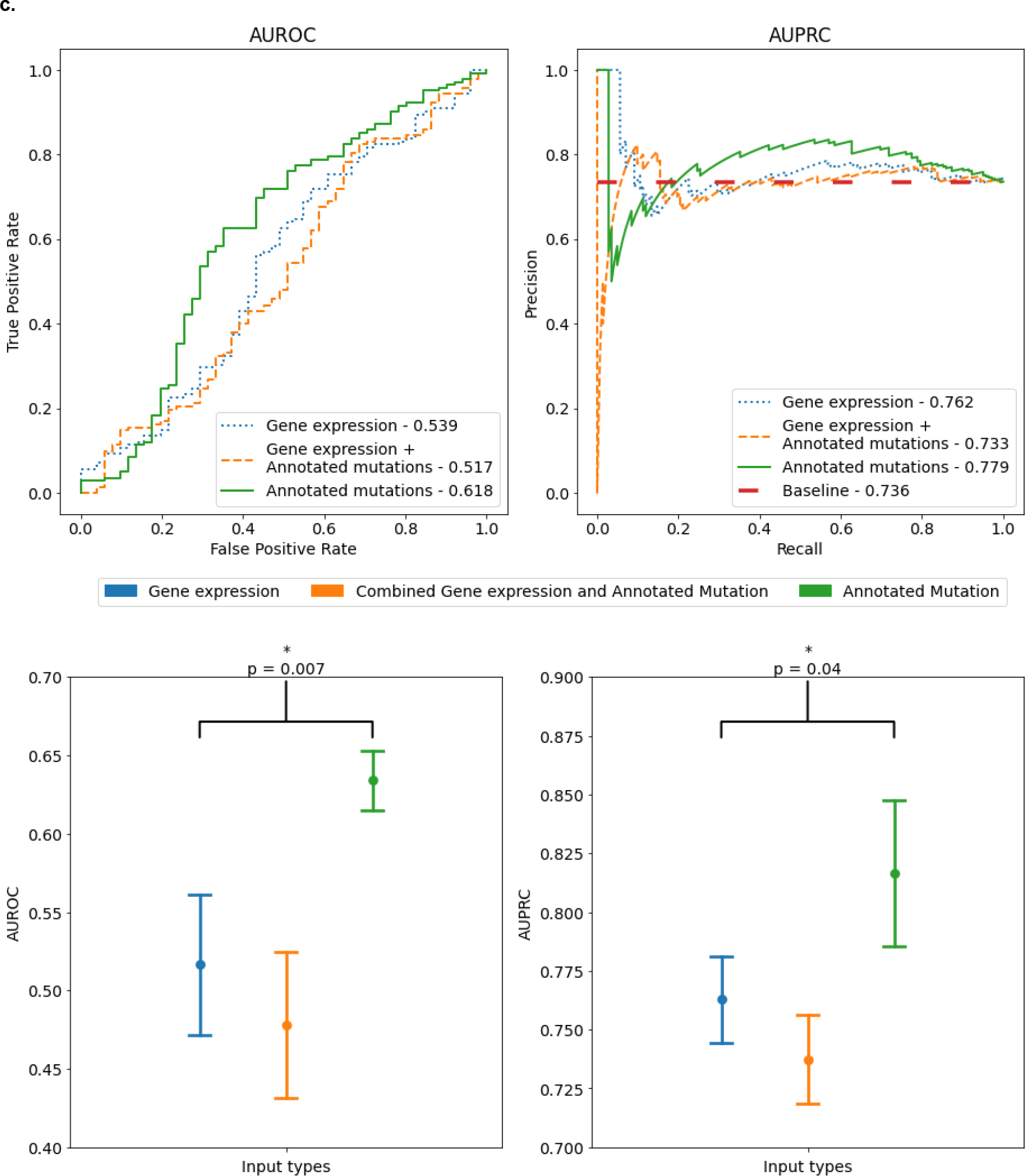
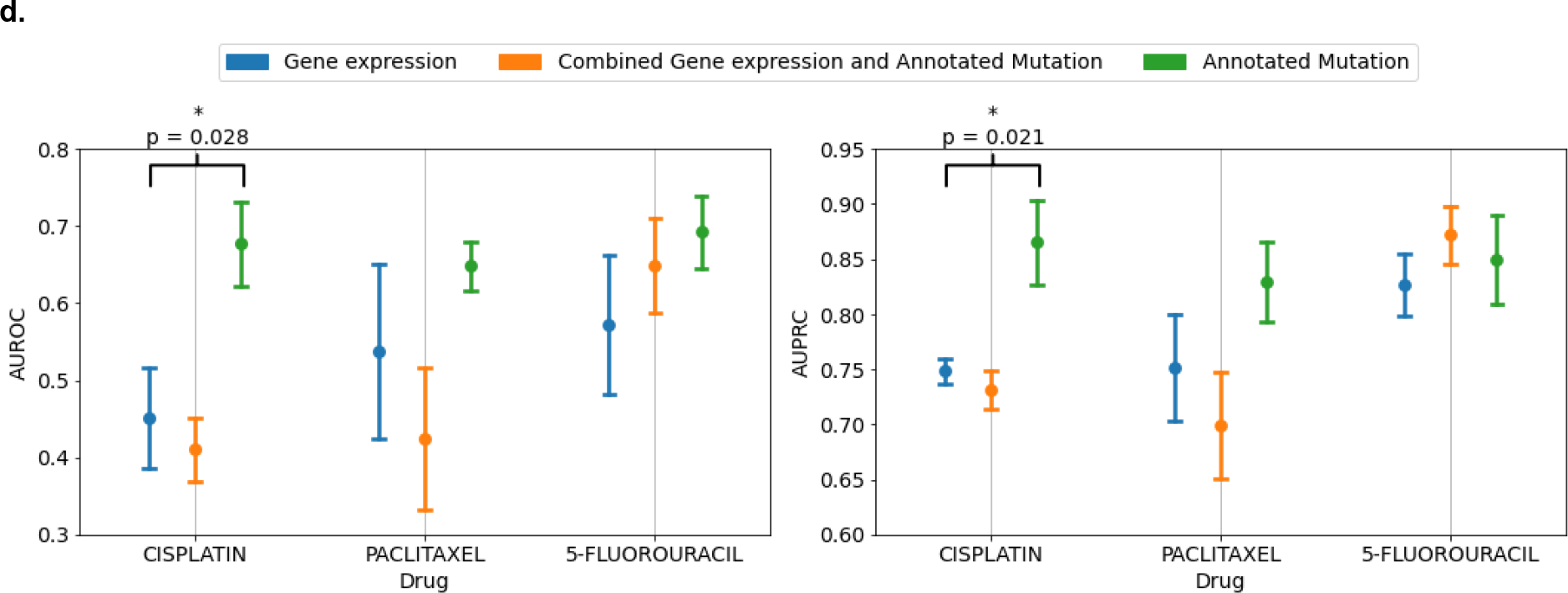
**(a):** Comparison of AUROC and AUPRC scores of response prediction for annotated mutations, copy number variations(CNV) and combination of the two. Performance was measured over 3 randomly chosen test splits (containing 90, 82 and 90 samples respectively). Top: Figure shows AUROC(left) and AUPRC(right) curves obtained after combining predictions on all 3 test splits. Bottom: Figure shows mean AUROC(left) and AUPRC(right) measured over 3 test splits with standard error. (**b):** Comparison of performance (mean AUROC, left and mean AUPRC, right) among annotated mutations, copy number variations(CNV) and combination of the two. Results shown separately for each drug in the data. **(c):** Comparison of AUROC and AUPRC scores of response prediction for annotated mutations, gene expression and combination of the two. Performance was measured over 3 randomly chosen test splits. Top: Figure shows AUROC(left) and AUPRC(right) curves obtained after combining predictions on all 3 test splits (containing 83, 80 and 84 samples respectively). Bottom: Figure shows mean AUROC(left) and AUPRC(right) measured over 3 test splits with standard error. Performance with annotated mutations was found to be significantly better than the other two input types. **(d):** Comparison of performance (AUROC, left and AUPRC, right) among annotated mutations, gene expression and combination of the two. Results shown separately for each drug in the data. * indicates statistical significance using a t-test between best and second-best performing inputs (*p* < 0.05).

Figure 3(b) shows the performance, across the three input data types, separately for cisplatin, paclitaxel and 5-Fluorouracil. The mean AUROC and AUPRC of DruID across the three agents are consistently higher when using annotated mutations alone as input, compared to CNV alone or CNV and annotated mutations combined. This reached statistical significance with AUPRC (*p*=0.019) and AUROC (*p*=0.004) for 5-Fluorouracil, and for AUROC for Paclitaxel (*p*=0.009).

Previous works have reported that gene expression has higher predictive value compared to mutation data (Partin 2023); however, transcriptomic data is not available in cNGS panel reports. We analysed a subset of patients from TCGA with both transcriptomic and genomic data available to compare the performance of DruID on 3 different input types: annotated mutations only, gene expression only and combined annotated mutations and gene expression. In all 3 cases, 324 genes represented in the FoundationOne CDx panel were used and performance measured over 3 test splits.

Figure 3 (c) shows the performance in terms of mean AUROC and AUPRC across the 3 input types over 3 test splits. Mutation data yields the best performance, in terms of mean AUROC (p=0.007) and AUPRC (p=0.040). Figure 3 (d) shows the AUROC and AUPRC values, across input data types by drug. The performance of DruID with mutational information alone was consistent across the three compounds and significantly better than gene expression containing inputs for cisplatin on both AUROC (*p*=0.028) and AUPRC (*p*=0.021). Differences between inputs for paclitaxel and 5-FU were non-significant.

We note that the performance of DruID, using annotated mutations, is comparable across all three drugs. However, when CNV, gene expression or their combinations with mutations were used, the performance varied across the drugs. In our experiments we consistently found that mutations with variant annotations yielded higher predictive signals.

### Validation of DruID on real world clinical datasets

We undertook cancer-specific clinical validation of DruID in two tumour types; colorectal (CRC), and ovarian cancer (OV). Data was collected from a single tertiary hospital (National University Hospital, Singapore) as part of an ongoing clinical study (Clinicaltrials.gov ID: NCT02078544). Patients enrolled underwent somatic NGS of tumour tissue or blood and treatment outcomes were recorded. We included those patients sequenced via FoundationOne CDx, utilising mutational information from the cNGS report (pathogenic and VUS). In light of our results above, showing worse performance with addition of CNV data, we did not incorporate CNV for the model training and evaluation.

For each analysis, we divided the respective datasets (CRC, OV) into train and test splits. DruID was trained on the patient train splits (CRC or OV) and cell line datasets, and was evaluated on the patient test split (CRC or OV). We evaluated the model ability to distinguish responders from non-responders ([PR or CR] versus [SD or PD] by RECISTv1.1 criteria). These analyses were done separately and are presented below.

#### Cancer-specific validation

The CRC dataset includes response to 3 drugs (5-FU, irinotecan, and oxaliplatin), in the first line metastatic settings. For the OV dataset, we included patients with advanced ovarian cancer with evaluable first line chemotherapy response (carboplatin/cisplatin, and paclitaxel).

Results of the performance analysis are shown in Fig. 4(a) with AUROC for each individual drug remaining above baseline of 0.5, with irinotecan most promising with AUROC=0.759 in CRC dataset and paclitaxel with AUROC=0.638 in OV dataset. Analysis of AUPRC(Figure 4(b) and 4(c)) highlights that DruID performs above baseline for the drugs considered.

**Figure. 4.**
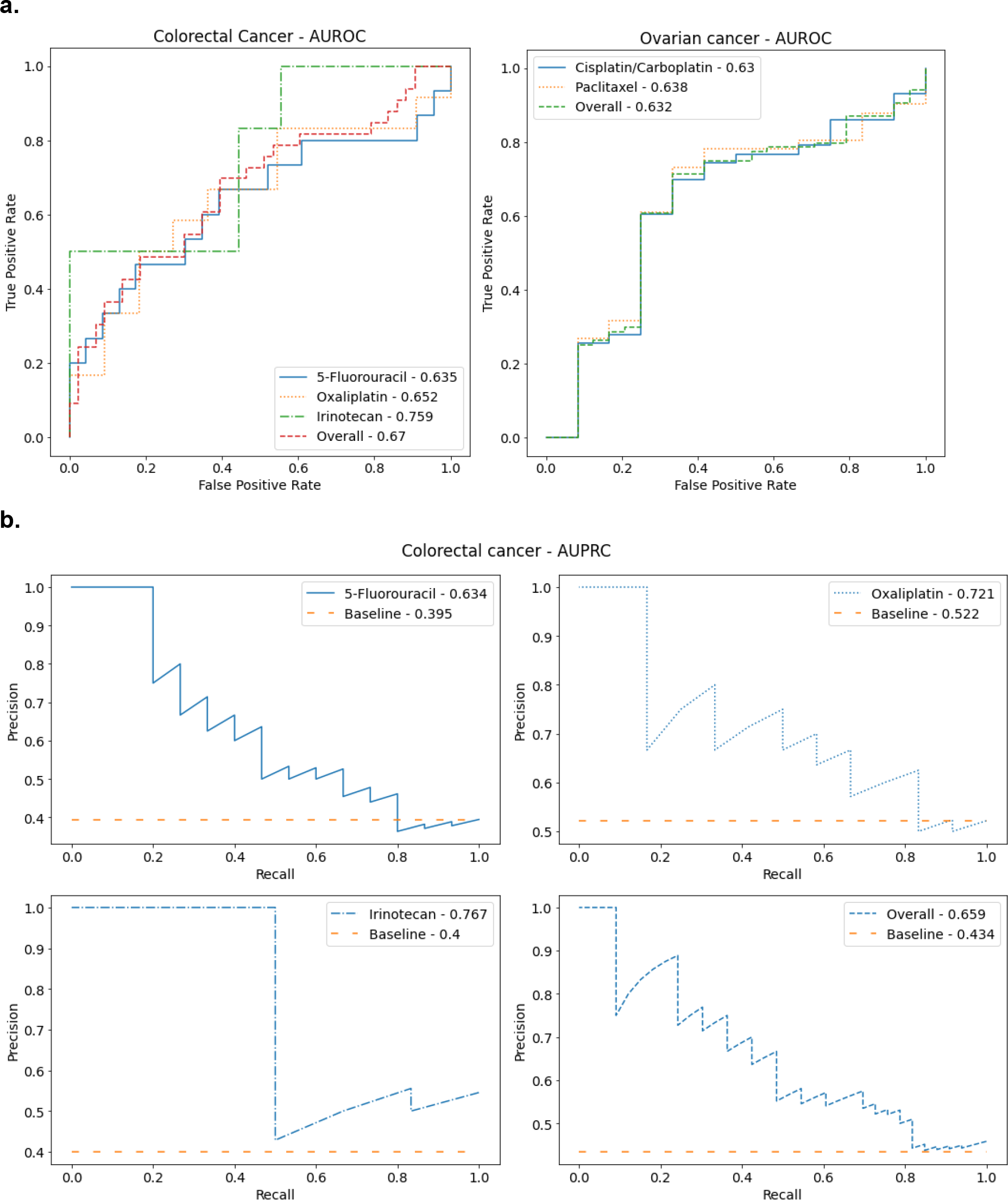
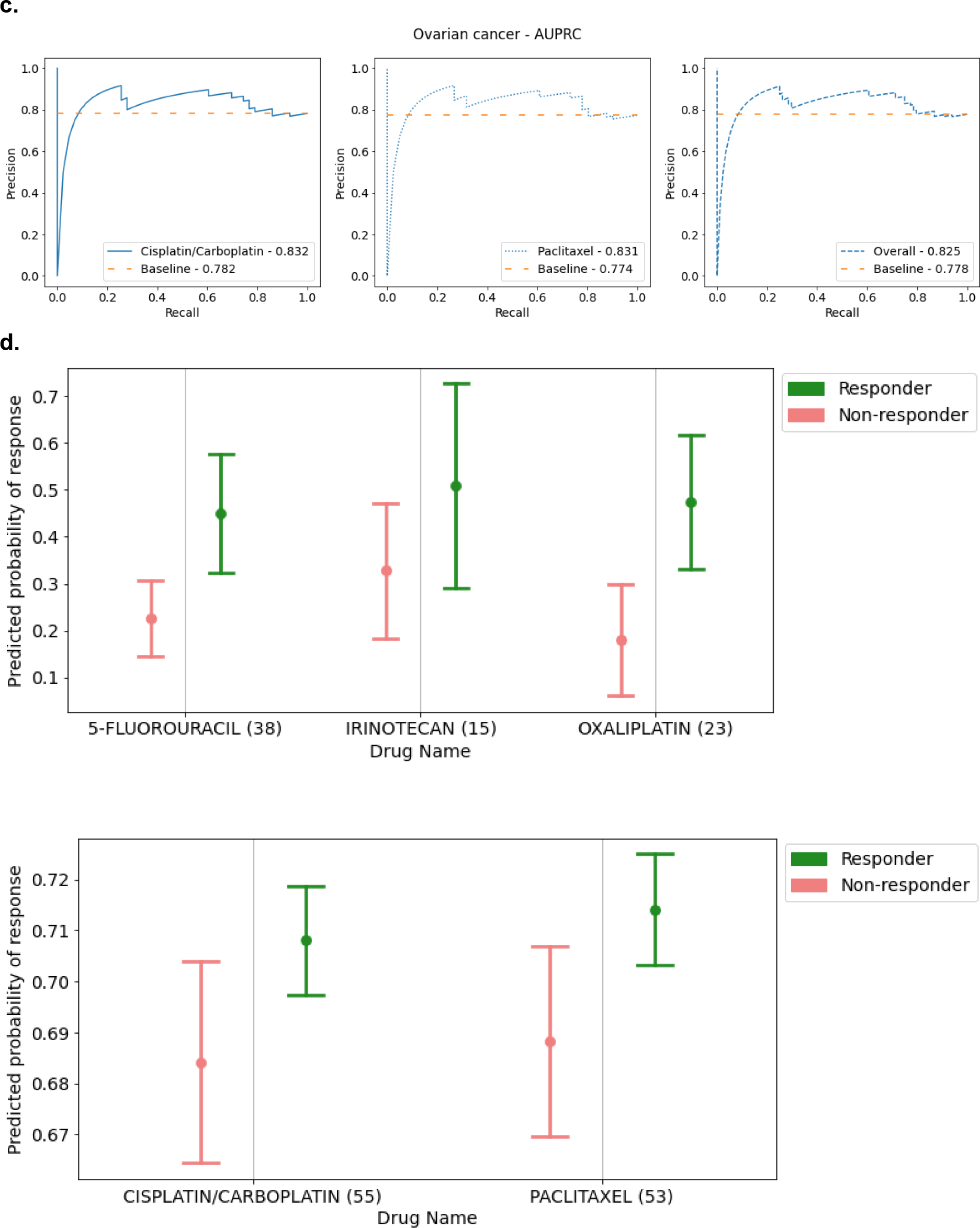
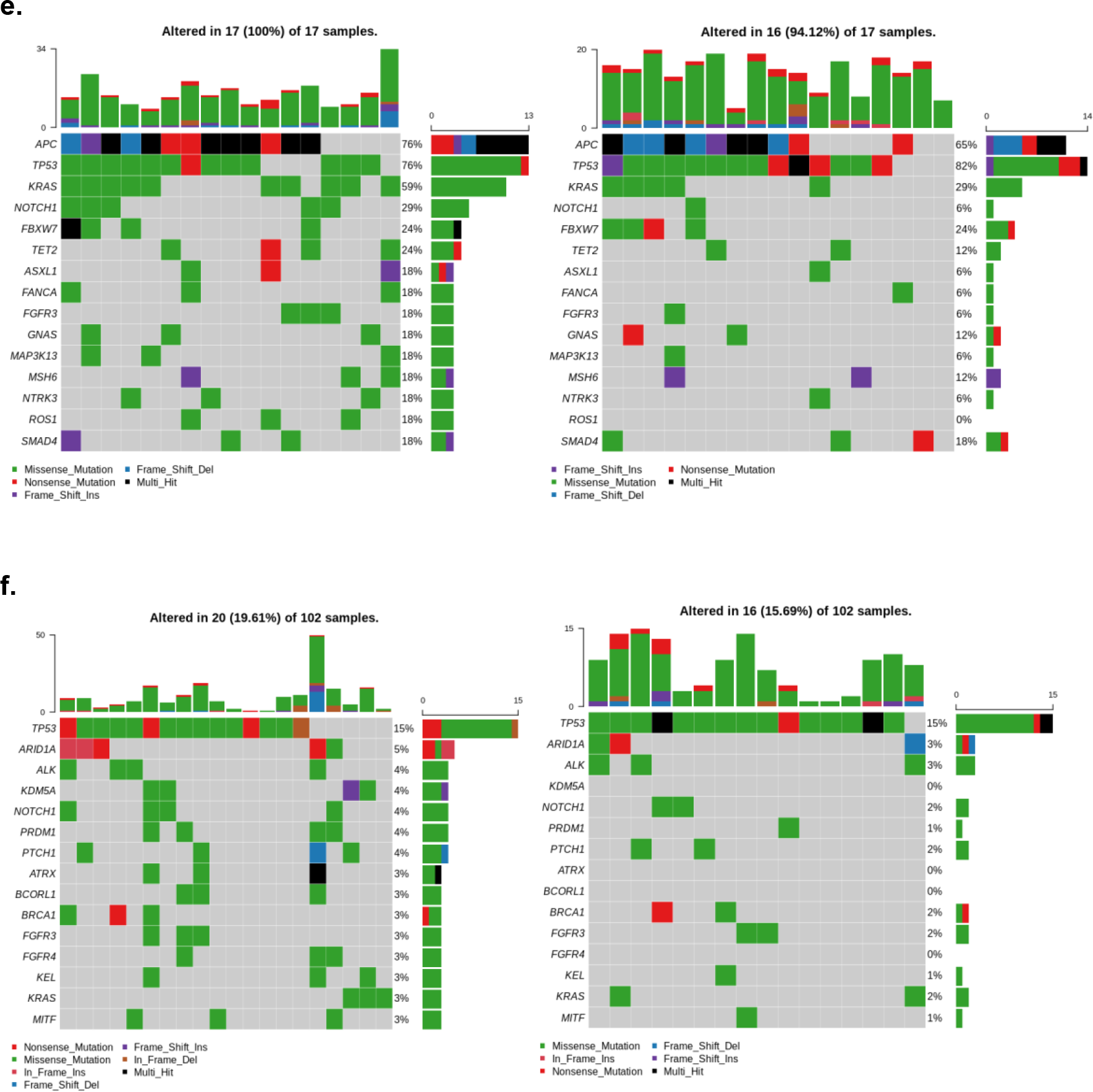
**(a)** Comparison of AUROC across 5-Fluorouracil, irinotecan and oxaliplatin in first line treatment of a cohort of stage IV colorectal cancer patients (left) and AUROC across Cisplatin/Carboplatin and Paclitaxel in first line non-surgical cohort of ovarian cancer patients (right). **(b)**: Comparison of drug specific AUPRC across 5-Fluorouracil, irinotecan and oxaliplatin in first line treatment of a cohort of stage IV colorectal cancer patients. Overall AUPRC is obtained across all 3 drugs. AUPRC for each drug is better than the corresponding baseline. **(c)**: Comparison of drug specific AUPRC across Cisplatin/Carboplatin and Paclitaxel in first line treatment of a cohort of ovarian cancer patients. Overall AUPRC is obtained across both drugs. AUPRC for each drug is better than the corresponding baseline. **(d):** Comparison of mean predicted probability of response, on true responders and non-responders in a cohort of first line stage IV colorectal cancer patients(top) and first line ovarian cancer patients (bottom). The plots indicate the mean±SEM DruID predicted probability of response (complete or partial response), for true responders and non-responders. **(e)**: Oncoplots showing frequent alterations in advanced(stage IV) colorectal cancer patients (IMAC dataset) based on DruID predicted response to 5-fluorouracil. i. Oncoplot(left) shows 15 most frequently altered genes in patients with predicted response in the bottom 20th percentile. Oncoplot(right) shows frequency of alterations in the same 15 genes listed in 4e(i) in patients with predicted response in the top 20th percentile. **(f):** Oncoplots showing frequent alterations in ovarian cancer patients (IMAC-GO dataset) based on DruID predicted response to Cis/Carboplatin. i. Oncoplot(left) shows 15 most frequently altered genes in patients with predicted response in the top 20th percentile. Oncoplot(right) shows frequency of alterations in the same 15 genes listed in 4f(i) in patients with predicted response in the bottom 20th percentile.

In comparing DruID’s predicted probability of response (CR or PR) for true responders versus non-responders, we see mean output values trend higher for true responders (Figure 4(d)), for CRC and OV cancer patient cohorts.

To assess for obvious discriminating features between DruID predictions we generated oncoplots (Fig. 4[e-f]) for cases with DruID predictions ranked in the bottom 20th percentile versus top 20th percentile for specific drugs in the validation datasets. Figure 4(e) shows oncoplots for 5-FU predictions in patients from the IMAC dataset. Mutations in KRAS (59% vs 29%) and NOTCH1 (29% vs 6%) are more frequent patients with a low predicted probability of response.

Oncoplots comparing gene alteration frequency in cases with top versus bottom 20th percentile of DruID response predictions to cis/carbo-platin across train and test splits of the IMAC-GO (OV) dataset are presented in Fig. 4(f). Low frequency of alterations limit interpretation. Alterations in *KDM5A* were seen in 4/17 (24%) cases with high predicted probability of response to cis/carbo-platin but no cases with low predicted probability.

## Discussion

Prior DRP methods that perform transfer learning have largely relied on gene expression data and WES panels. However, this data is unavailable in a clinical setting, where often, only a subset of recurrently altered genes are sequenced using cNGS, to identify mutations and copy number variations. Through our empirical evaluation (Fig. 1), we have shown that state-of -the-art DRPs can perform comparably with mutational information from a cNGS panel and whole exome sequencing. This is of significance, potentially increasing the number of patients for which a DRP such as DruID could be utilised, as cNGS is increasingly being undertaken as a standard of care in oncology practice.

However, due to the relatively poor performance of existing methods on cNGS inputs, we propose a novel drug response prediction model(DruID) that handles two key challenges arising in the clinical context namely (1) sparse nature of mutation data and (2) limited availability of patient drug response data. While most methods handle the distributional differences between cell lines and patient genomic profiles, most methods do not handle the differences in drug response measurements across the two domains (Table 2). Further, most of these methods were trained and evaluated on gene expression data rather than mutation data. To the best of our knowledge, DruID is the first model that uses variant annotations for mutation data processing. Similar to prior methods(Velodrome and AITL), DruID simultaneously handles distribution differences in the mutation profiles and differences in the way drug response is measured across cell lines and patients. Unlike most of the prior methods, DruID has the capability to utilise unlabelled patient data. For further details, see Supplementary Section “Background on other ML approaches”.

Our results show that DruID outperforms other state-of-the-art DRP methods (Fig. 2) on publicly available TCGA data. DruID shows robust performance on two clinical cohorts of colorectal and ovarian cancer patients (Fig. 4). These tumour types have widely different biology and molecular profiles (Dienstmann 2014, Haunschild 2021) highlighting DruID’s performance is not dependent upon the presence of certain gene/mutation signatures that may be specific to one tumour. Validation on other patient cohorts can further establish this generalisability. A limitation of our current validation in the TCGA and IMAC/IMAC-GO datasets is the modest number of patients and drugs included, due to restricted availability of labelled response data. This is a problem encountered in the validation of many DRPs, with the acquisition of reliable patient response data key to model training and performance.

The use of CNV and gene expression data did not improve results when compared to those obtained from annotated mutations alone (Fig. 3). This is contradictory to prior work on cell lines, where gene expression data showed the best performance(Partin 2023). The findings in our experiment suggest that DruID is superior in its ability to handle sparse mutation data; however our test set sizes are quite small, these results may not generalise to all patient cohorts and would need to be validated on larger patient datasets. In our experiments, we also find that the performance with combinations of different data types (mutations and CNV; mutations and gene expression) was found to be lower than that of each of the individual data types (CNV and gene expression respectively), in most cases. This suggests that the modelling approach can be further improved with respect to integrating diverse data types. One approach could be to use multimodal techniques to handle different data types.

Assessment of mutational profiles of patients with low versus high predicted probability of response suggests DruIDs ability to identify biomarkers of poor response/prognosis consistent with prior knowledge. Alterations in *KRAS* and *NOTCH1* appeared more frequently in predicted non-responders from the CRC patient dataset (Fig 4[e]). This is consistent with the known function of *KRAS* alterations as a poor prognostic marker in colorectal cancer (Zhu 2021). The role of *NOTCH1* as a prognostic marker is not as clear in CRC, but in oesophageal SCC it is reported to be associated with cancer progression and lower response rates. (Song 2015, Jackstadt 2019). In the analysis of ovarian cancer patients’ mutational profile by predicted probability of response, it is difficult to draw strong patterns from the frequencies presented (Fig. 4[f]). A possible trend is seen for *KDM5A* alterations, appearing in patients with high predicted probability of response but absent in those with low predicted probability. This will benefit from further patient analysis. The role of KDM5A in cancer is continuing to be elucidated, but overexpression is thought to drive progression(Ren 2020).

The current design of our model could be improved further. While patients often undergo a treatment regimen comprising multiple drugs, in our experiments we treat each drug as being administered independent of the others. To consider a regimen as a whole, a possible approach could be to use the combination of drugs as an input to the drug embedder network. The multi-task learning architecture in Stage III of DruID also allows the addition of related tasks. Patient survival information can be incorporated into the model in the form of an additional task. Further improvements can be made to the model’s explainability as well. Currently, it is not inherently explainable. Explainable algorithms (Jiménez-Luna 2020) can be used over the model predictions to obtain useful insights to improve user confidence and guide clinical decision making. While in Stage II, DruID leverages available unlabelled data, Stage III relies on labelled patient data similar to earlier DRP methods.

DruID has significant potential as a clinically applicable DRP. Due to its design it can be fine-tuned for any drug or cancer type provided sufficient training data (both unlabelled and labelled) is available. Additionally, it has the possibility to be utilised as a drug repurposing tool as it can provide response predictions for previously unseen drugs by utilising drug molecular information as a model input (Fig. 2). This is of potential significance to patients with refractory advanced malignancies, who in the absence of an actionable mutation being identified on cNGS, will often undergo empiric anti-cancer therapy with low expected response rates. The prospect of a drug repurposing tool that can utilise cNGS data to give a personalised treatment recommendation based on a tumour mutational profile is both exciting and appealing. Such an application of a DRP model requires prospective validation, the first steps of which we are undertaking in an ongoing trial incorporating DRP recommended therapy in patients with refractory solid organ malignancies in Singapore (NCT05719428).

## Conclusion

In this paper, we evaluated state-of-the-art DRP models on the limited subset of genes sequenced in cNGS panels and established that gene panels can perform as well as WES panels for DRP. To improve the performance of DRP models on cNGS panels, we present a novel transfer learning based DRP algorithm - DruID - which can handle sparse nature of cNGS mutation data and the limited availability of patient response to drugs. Results presented show DruID to be superior to existing state-of-the-art DRP methods on a pan-cancer TCGA dataset with satisfactory performance seen on two cancer-specific clinical datasets. While we have utilised a panel of genes specific to one commercially available cNGS test, DruID can be altered to work on any panel gene set. Future work developing such tools for drug repurposing endeavours may provide further clinical applications.

## Methods

### DruID Method Description

In the following sections, we describe each of the three stages of DruID(Fig. 2a) in more detail.

#### Stage I: Variant Annotations

For each point mutation in the input dataset, we generate annotations using the following tools:

1. ClinVar (Landrum 2017) – provides clinical significance of each mutation. We group these together into 3 broad categories - pathogenic, benign and variants of unknown significance (VUS).
2. GPD (Li 2020) – provides annotations based on the location of each mutation - protein information unit (PIU), linker unit (LU) and non-coding unit (NCU).
3. Annovar (Wang 2010) – provides annotations for each mutation that indicates if it is deleterious or not, from 17 different prediction algorithms (as shown in Supplementary Table 3). These are aggregated (via mean) to calculate a d-score for each mutation.

The categories from GPD, Clinvar and the score from Annovar are shown on the top right of Fig. 5. These are aggregated to obtain gene-level features, shown on the bottom right of Fig. 5. Below we describe more details.

**Figure 5:**
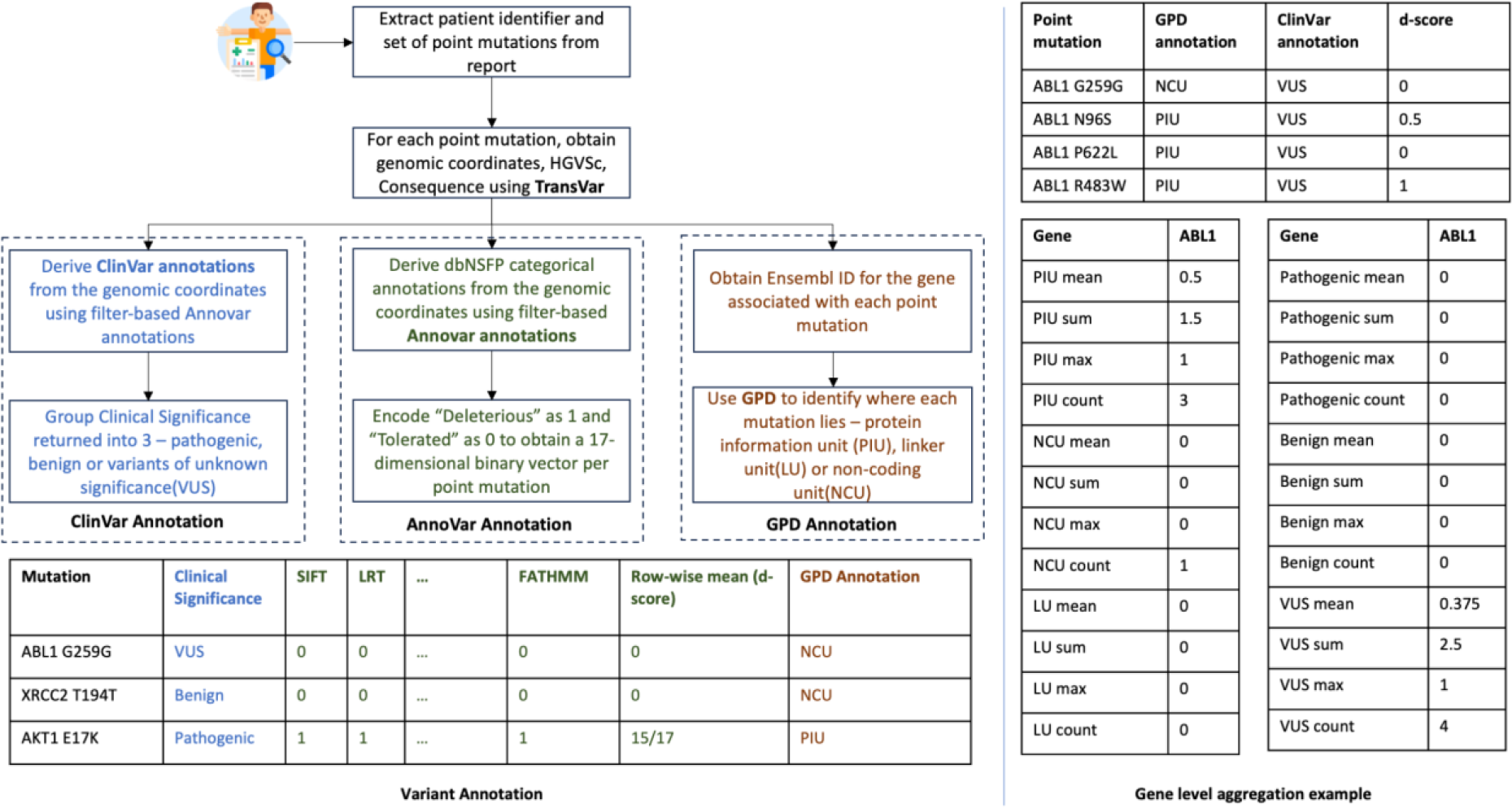
Stage (I) Variant Annotation starts with the extraction of mutations from the cNGS report. We first use the protein level annotation feature of TransVar on the extracted mutations. This returns the genomic coordinates, consequence attributes for the mutation. The output of TransVar, specifically the genomic coordinate information, is used to obtain both ClinVar and Annovar annotations. ClinVar annotations, indicating the clinical significance of the mutations, are generated using filter-based Annovar annotations with ClinVar database. The resulting annotations are grouped into one of 3 categories - pathogenic, benign and variants of unknown significance. Annovar annotations indicate whether a mutation is deleterious or tolerated, using the predictions from 17 algorithms. These annotations are encoded as a binary vector and mean aggregated to obtain a d-score per mutation. GPD annotation needs an Ensembl ID for each gene, which is generated using the MyGene package. GPD annotates each mutation based on its location as lying in a protein information unit(PIU), linker unit(LU) or non-coding unit(NCU). The output of the annotation is shown in the table on the bottom left. These are further aggregated at a gene level, as indicated on the right. All mutations in a gene belonging to each of the 3 GPD categories are aggregated using mean, max, sum and count. This is repeated for each of the 3 ClinVar categories to obtain 4 features per category. This results in 24 features for each gene. Thus for the Foundation One report comprising 324 genes, a 324 * 24 = 7776 dimensional annotated mutation vector is constructed per patient.

##### Processing for ClinVar, GPD and Annovar

Before annotations from ClinVar (Landrum 2017), Annovar (Wang 2010) and GPD (Li 2020) can be obtained, we use TransVar (Zhou 2015), which takes as input a point mutation and provides the location of the mutation on the genome. The TransVar output is used by Annovar, GPD and ClinVar.

For GPD, the input is expected to be in the MAF(Mutation Annotation File) format. The TCGA dataset is directly available in this format. However, for the NUH ovarian cancer and colorectal cancer datasets, an additional processing step must be done to obtain the Variant Classification attribute. We use the Consequence field returned by TransVar, to obtain this attribute. The Consequence field indicates if the input point mutation is a missense, synonymous, nonsense, frameshift or splice site mutation. GPD also needs the Ensembl Gene ID, which we generate from Entrez Gene ID using myGene python package (https://docs.mygene.info/projects/mygene-py/en/latest/). GPD returns a category (from PIU, LU and NCU) for each point mutation based on its location.

For each point mutation, Annovar returns 17 categorical scores, one score for each of the algorithms listed in Supplementary Table 3. We convert these 17 categorical values into a single score, called d-score, as follows: if *x* of the 17 algorithms flag a point mutation as deleterious, we set the d-score of the point mutation to be *x*/17. Thus after Annovar annotation, each point mutation has a d-score, whose value lies between 0 and 1. ClinVar annotation provides a clinical significance for each point mutation. Each point mutation is annotated as pathogenic, benign or as a variant of unknown significance(VUS). The mapping from ClinVar generated annotation to these 3 categories is available in Supplementary Table 4.

##### Gene-level Features

After obtaining annotations from ClinVar, Annovar and GPD, we aggregate the d-scores of the point mutations at a gene level. As shown in Fig. 5, a gene can have multiple point mutations, and each point mutation can belong to one of 3 ClinVar categories and one of 3 GPD categories. Each point mutation is also associated with a d-score. To aggregate these at a gene level, we obtain the mean, max, sum and count of point mutations present in each gene, in each of the ClinVar and GPD categories. For example, from Fig. 5, we see that the 4 point mutations in the ABL1 gene are distributed across PIU and NCU GPD categories and VUS ClinVar category. We can aggregate across all ABL1 mutations in the PIU and NCU categories, as well as all ABL1 mutations in the VUS category separately. Each gene now has 6 subcategories (GPD - PIU, LU, NCU and ClinVar - pathogenic, benign,VUS), each with 4 statistics (mean, max, sum, count), resulting in 24 features per gene per patient sample. If a gene has no mutations, it is represented as a 24-dimensional zero vector.

#### Stage II: Unsupervised domain-invariant representation learning

In this stage, we use Variational Autoencoders (VAE), which are unsupervised generative neural models (Kingma and Welling 2013), to obtain domain-invariant low-dimensional representations of the input data. One VAE is used for each domain (cell lines, patients) as shown in Fig. 6. The VAEs are trained to maximise the likelihood of the data assuming zero-inflated data distributions which allows us to model both count and real-valued data (through Zero-inflated Negative Binomial and Zero-inflated Normal distributions respectively) and with varying levels of noise and sparsity (Eraslan 2019, Mariappan 2022). To ensure that a shared embedding space is learnt across the two domains, an alignment loss is introduced during training of the VAEs. This is achieved through the CORAL loss (Sun 2016). Note that this stage does not need any drug response data from either domains and is done in an unsupervised manner. Doing so allows effective use of any patient or cell line data that has just genomic data, unlike previous approaches such as AITL, TCRP, PACE. We now formally describe the details.

**Figure 6:**
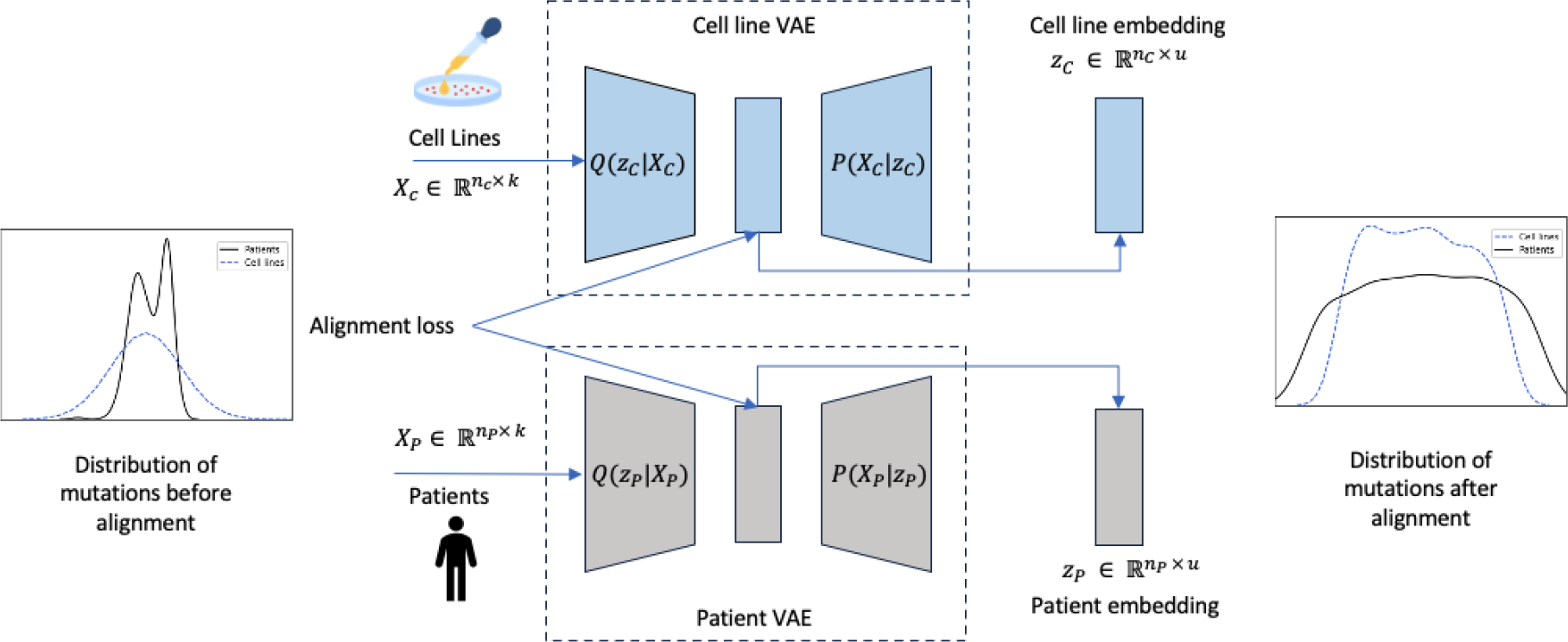
Stage (II) Unsupervised domain-invariant representation learning involves the use of two separate variational autoencoders, one per domain (cell line or patient). The VAEs take as input the annotated mutation vectors generated in Stage (I) Variant Annotations and learn a lower dimensional representation for each domain. To account for the sparse nature of the input data, the VAEs are trained to maximise the likelihood of the data following a zero-inflated distribution(zero-inflated negative binomial for count data and zero-inflated normal for real-valued data). To ensure the domain-invariant nature of representations, an alignment loss (CORAL loss) is introduced between the representations learnt from both VAEs. This stage does not require labelled samples and can be trained in a fully unsupervised manner.

Let 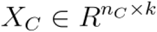 and 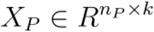 denote the input data associated with cell lines and patients respectively, where *n*_*C*_ denotes the number of cell lines, *n*_*P*_ denotes number of patients and *k* denotes number of input features. In the VAEs used to obtain representations *z*_*C*_ and *z*_*P*_ for cell lines and patients respectively, the probabilistic encoders *E*_*C*_ and *E*_*P*_ - which learn distributions *Q*(*z*_*C*_|*X*_*C*_) and *Q*(*z*_*P*_|*X*_*P*_) respectively - infer the mean *μ*_*C*_, *μ*_*P*_ and standard deviation *σ*_*C*_ and *σ*_*P*_ of the normal distributions of the latent variables. Thus, each input vector is mapped to a distribution and we use the inferred mean vectors as the latent representations *z*_*C*_ and *z*_*P*_ for downstream tasks. The probabilistic decoders *D*_*C*_ and *D*_*P*_ which learn distributions *P*(*z*_*C*_|*X*_*C*_) and *P*(*z*_*C*_|*X*_*C*_) respectively use zero-inflated distributions to model varying levels of sparsity in the reconstructed data. Let 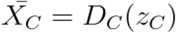 and 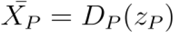 denote the reconstructed outputs from the decoders. These are used to learn the parameters of the zero-inflated distribution (Π Ω, and Θ), using linear layers followed by relevant activation functions. In the equations below, *e* can be *C* or *P* to denote cell lines or patients.

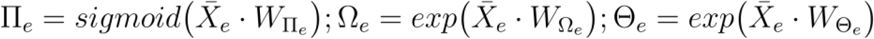

Each VAE is trained by minimising the negative log-likelihood of the data distribution and a KL divergence term (that acts like a regularizer). Thus, we have the combined loss term

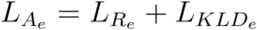

where

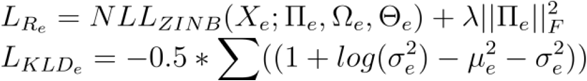

More details of the VAE construction and loss functions can be found in (Mariappan 2022).

The VAE architecture, as described above, can obtain low-dimensional dense representations of cell lines and patients. However, it does not ensure domain invariance in the representations. To achieve domain invariance, we use the CORAL loss(Sun 2016) to minimise the difference in covariance of the domain-specific representations. If *C*(*z*_*C*_) and *C*(*z*_*P*_) are the covariance matrices of cell lines and patients representations *z*_*C*_ and *z*_*P*_, the CORAL loss is defined as 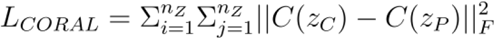, where 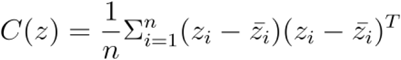. Here *n* is the number of samples in the batch, *n*_*Z*_ is the size of the dimensional space *Z* and 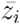 is the mean of the representations, across samples in the batch. Thus the overall loss function optimised in the pretraining stage is 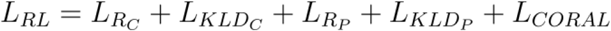. The hyperparameters used in this stage are listed in Table 3.

**Table 3:** Key hyperparameters used to train DruID.

| Stage II Unsupervised Domain Invariant Representation Learning |  |  |  |
| --- | --- | --- | --- |
| Hyperparameter | CCLE-TCGA | CCLE-CRC | CCLE-OV |
| VAE hidden layer dimensions (encoder and decoder) | No pre-training; uses model pre-trained on CCLE-IMACGO for fine tuning | [128, 64] | [128, 64] |
| VAE activation function |  | tanh | tanh |
| Learning Rate |  | 1e-5 | 1e-5 |
| Epochs |  | 1000 | 1000 |
| Convergence threshold |  | 1e-5 | 1e-5 |
| Stage III Multi-task Drug Response Prediction |  |  |  |
| Hyperparameter | CCLE-TCGA (fine-tuning) | CCLE-CRC | CCLE-OV |
| Batch size | 256 | 256 | 256 |
| Epochs | 50 | 500 | 500 |
| Cell line embedder learning rate | 1e-4 | 1e-4 | 1e-6 |
| Patient embedder learning rate | 1e-4 for 5-fu; 1e-3 for Cisplatin and Paclitaxel | 1e-3 | 1e-4 |
| AUDRC and RECIST predictor learning rate | 1e-3 for 5-fu; 1e-5 for Cisplatin; 1e-6 for Paclitaxel | 1e-6 | 1e-6 |
| Drug embedder learning rate | 1e-4 | 1e-4 | 1e-4 |

Table shows the key hyperparameters used to train DruID, specifically in stages II and III on train split 0 for each dataset used in the experiments. For CCLE-TCGA dataset, the initial pre-training is done on the CCLE and IMAC-GO datasets with further fine-tuning done per drug. In all other cases, initial pre-training and fine-tuning are done on the same dataset.

#### Stage III: Multi-task drug response prediction

Drug response in cell lines and patients are known to be different due to biological and environmental differences. Further, measurement of responses are different – real-valued AUDRC for cell lines and categorical RECIST scores for patients, which correspond to regression and classification tasks. To build a model that can learn from both domains and, at the same time, predict for each task separately we use multi-task learning (MTL), a well established paradigm for jointly learning models for multiple correlated tasks.

There are three inputs to the MTL model – a cell line representation, a patient representation and a drug representation. The first two are obtained from the encoders of the VAEs from stage II. To obtain a drug representation, we build another feedforward neural network, called the drug embedding network, which takes as input the drug’s binary Morgan fingerprint (Morgan 1965). Corresponding to each task, AUDRC prediction and RECIST prediction, we have a separate feedforward neural network. The output of the drug embedding network is concatenated separately with the cell line and patient representations from the respective encoders and passed through one of the two task-specific networks. The concatenated drug - cell line representation is passed through the AUDRC prediction network while the concatenated drug - patient representation is passed through the RECIST prediction network. Let 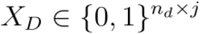 be the input features associated with the drug and *z*_*D*_ = *E*_*D*_(*X*_*D*_) be the drug embedding from the drug network *E*_*D*_. The input to the AUDRC predictor network *f*_*C*_ is *z*_*C*_ ⊕ *z*_*D*_ where ⊕ denotes concatenation. Likewise, the input to the RECIST predictor network *f*_*P*_ is *z*_*P*_ ⊕ *z*_*C*_. It is possible to ‘attach’ the VAE encoders from stage I, to further train them with the rest of the network in a supervised manner; in most cases, this is found to improve performance.

The entire network (Fig. 7) is trained using two objective functions – the MSE loss for AUDRC regression and the BCE logit loss for RECIST classification. Let *y*_*AUDRC*_ and *y*_*RECIST*_ denote the ground truth labels and 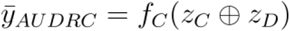 and 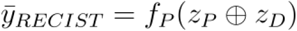, the BCE logit loss and MSE loss are calculated as follows

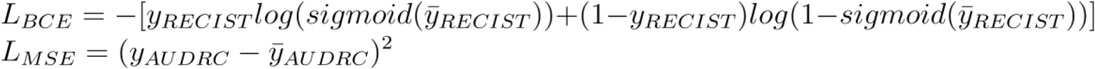

A common approach to train MTL models is by minimising the weighted sum of the losses for each task, where the weights specify relative priorities among the tasks; this is known as the linear scalarization approach. However, when the tasks are conflicting, it may not be possible to optimise all the objectives simultaneously and trade-offs between tasks may be required. In such cases, Pareto optimal solutions, obtained through multi-objective optimization, are natural choices where each optimal solution is non-dominated, i.e., no objective value can be improved further without degrading some other objectives. The efficacy of multi-objective optimization for MTL has been demonstrated in, e.g., (Sener & Koltun, 2018; Mahapatra and Rajan 2020).

**Figure 7:**
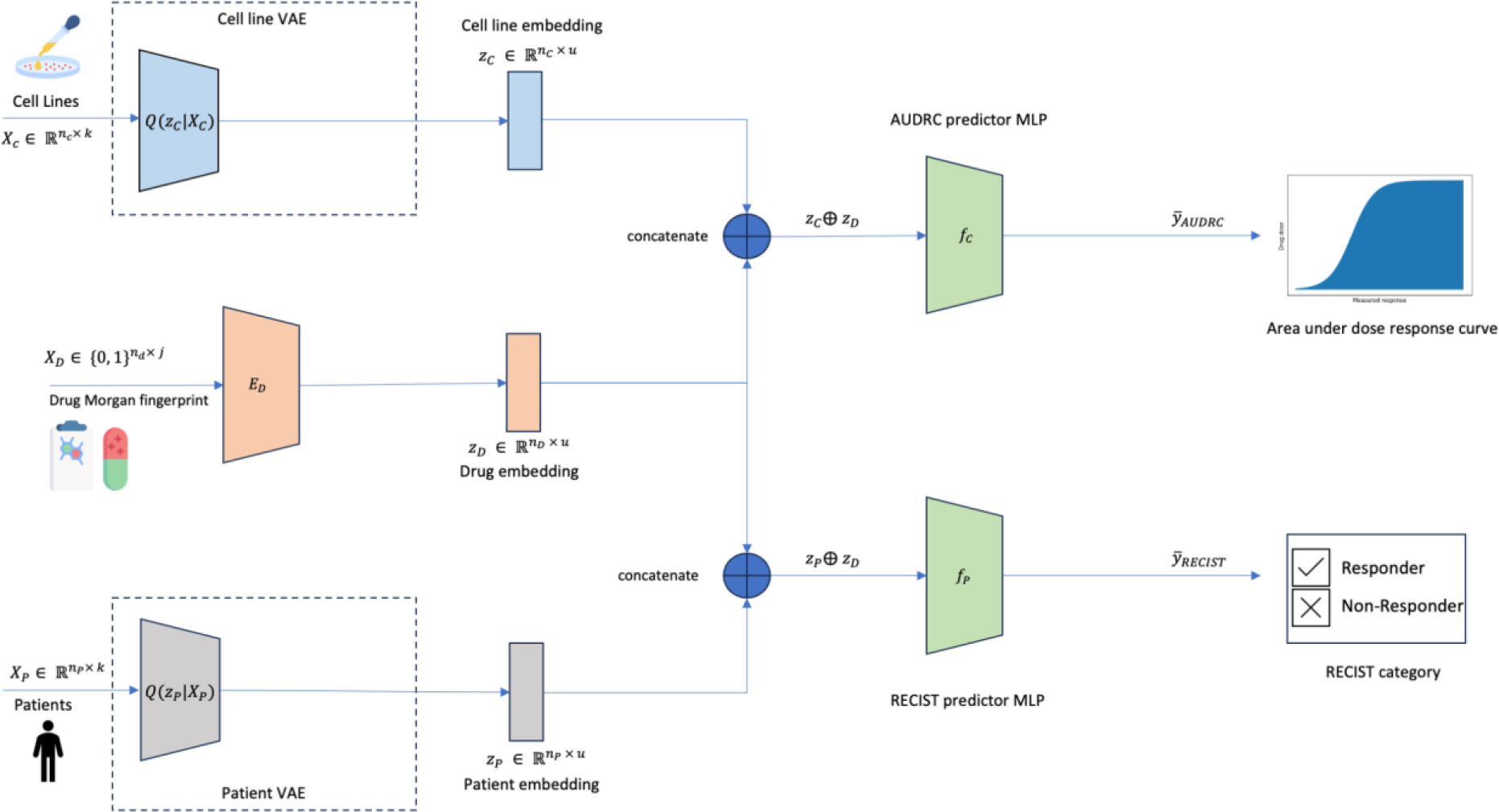
Stage (III) Multi-task drug response prediction uses the representations learnt for patients and cell lines from Stage (II) Unsupervised domain-invariant representation learning. This is achieved by attaching the trained encoders of the VAEs from Stage (II). In addition, at this stage, we also introduce drug information in the form of the Morgan fingerprint. This is passed through a feedforward neural network. This is followed by a pair of task-specific feedforward neural networks - one for the regression task for AUDRC prediction on cell lines and another for the classification task of RECIST category prediction on patients. The representations of cell lines are concatenated with the drug representation, before being passed through the AUDRC predictor multi-layer perceptron(MLP). Likewise, the patient representation is concatenated with the drug representation and fed into the RECIST predictor multi-layer perceptron(MLP). Binary cross-entropy loss and mean square error are calculated for the classification and regression task respectively. The network is trained for the two tasks through multi-objective optimization (Chebyshev scalarization).

There can be multiple (possibly infinite) Pareto optimal solutions, represented by the Pareto front, each solution with a different trade-off between the conflicting objectives. For non-convex objective functions common in machine learning, linear scalarization cannot guarantee reaching every possible solution on the Pareto front (Miettinen 2004, Lin 2019, Boyd 2021). This can be guaranteed through the use of Chebyshev scalarization (Van Moffaert 2013) that we utilise for training our MTL network: *L*_*MTL*_ = *max*(*λ*_*P*_*L*_*BCE*_, *λ*_*C*_*L*_*MSE*_) where *λ*_*P*_ and *λ*_*C*_ are hyperparameters denoting the weights assigned to each loss term. Details of hyperparameters are listed in Table 3.

#### Inference

The trained network can be used to predict drug response for a given mutation profile and drug. Note that only the drug embedding network, patient-specific VAE encoder and RECIST predictor networks are required during inference as shown in Fig. 8.

**Figure 8:**
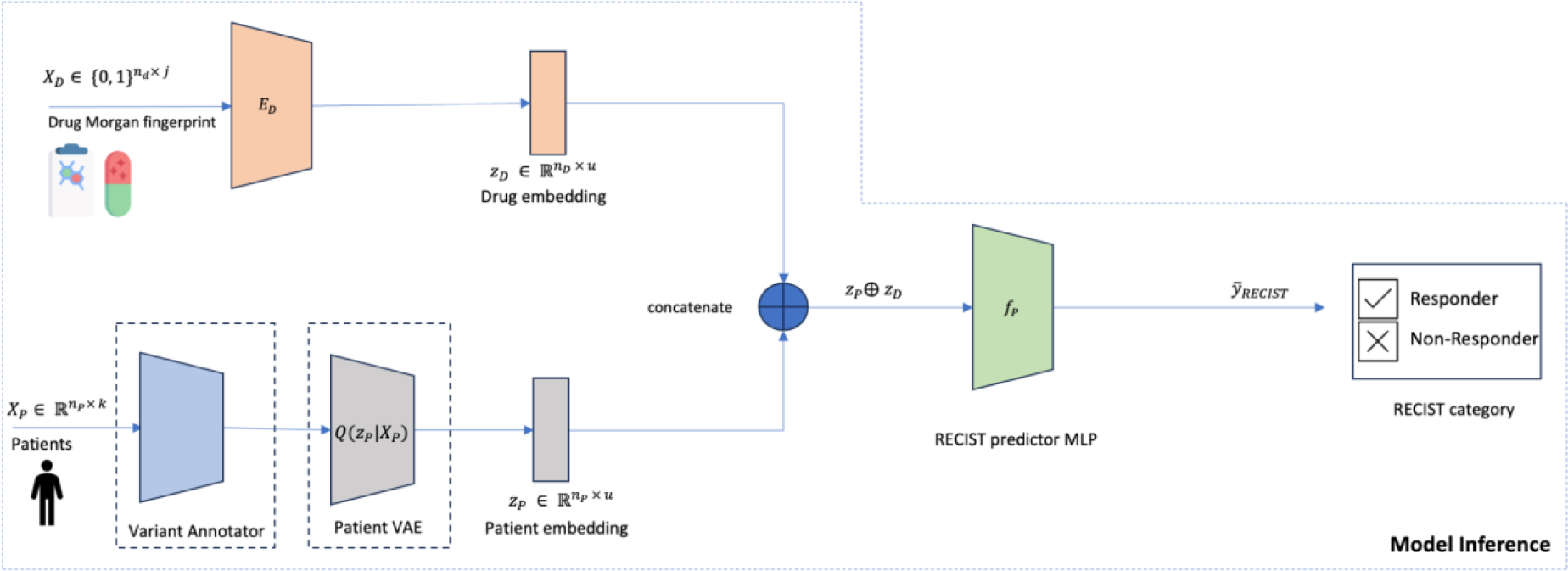
During inference, a (patient, drug) pair is passed in. The trained network takes as input the patient mutations and annotates it using Stage (I) Variant Annotations. This is passed through the trained encoder of the patient VAE to obtain the patient representation. The Morgan fingerprint of the drug in the test pair is passed through the trained drug embedder. The drug and patient representations are concatenated and passed through the RECIST predictor network, which returns a predicted probability of response (RECIST category of CR or PR).

#### Comparison of DruID with previous modelling approaches

The architecture proposed above for DruID allows us to utilise unlabeled patient data, while simultaneously modelling both the distributional differences in omics inputs and differences in drug response measured across patients and cell lines. DruID also handles sparse nature of mutation data and enables the use of variant level information associated with these mutations. While some of these can be handled by extant DRP methods, DruID can perform all of these simultaneously. Table 2 shows a comparison of DruID against extant DRP methods.

### Datasets

#### TCGA

The raw mutation data was obtained from The Cancer Genome Atlas (Weinstein 2013) GDC portal (https://portal.gdc.cancer.gov/). The response data was obtained from (Jia 2021). We only consider the cancer types belonging to the following TCGA projects/cancer types - LUAD, STAD, HNSC, SKCM, BLCA, UCEC, COAD, LUSC, BRCA, CESC. We further retain the TCGA samples which have a corresponding RECIST v1.1 response to the drugs Cisplatin, Paclitaxel, 5 - Fluorouracil, Gemcitabine, Docetaxel, Cyclophosphamide. These are the drugs with at least 50 TCGA samples having a documented RECIST response. We further convert the RECIST labels into two categories - complete response(CR) and partial response(PR) are grouped together as responders while stable disease(SD) and progressive disease(PD) are grouped together as non-responders.

#### Mutations

We first filter out and retain those TCGA samples which have a mutation classified as one of “Missense_Mutation”, “In_Frame_Del”, “Splice_Site”, “Nonsense_Mutation”, “Frame_Shift_Ins”, “Frame_Shift_Del”, “Nonstop_Mutation”, “Translation_Start_Site”, “In_Frame_Ins”.

#### Gene Expression

The gene expression data from the TCGA GDC portal(v1.29.0) (Weinstein 2013) was used directly.

#### IMAC Colorectal Cancer (IMAC-CRC)

The raw data was obtained from patients with advanced colorectal cancer enrolled and consented into the Integrated Molecular Analysis of Cancer (IMAC) study. The IMAC study is an ongoing prospective trial using broad panel sequencing of refractory solid-organ malignancies to identify targetable molecular alterations in the Phase I unit of the National University Cancer Institute, Singapore (NCIS). We retained patients with successful sequencing on FoundationOne CDx and available response data from their first line therapy in the metastatic setting. Drugs with more than 10 response events (patient, drug pairs) and available smiles string were included in subsequent training. These included 5-fluorouracil (includes capecitabine), irinotecan, oxaliplatin and cetuximab. We converted the RECIST labels into two categories - complete response(CR) and partial response(PR) are grouped together as responders while stable disease(SD) and progressive disease(PD) are grouped together as non-responders.

#### IMAC-GO Ovarian Cancer (IMAC-OV)

The raw data was obtained from patients with advanced ovarian cancer enrolled and consented into IMAC-Gynaecologic Oncology (IMAC-GO) study, a prospective study using broad panel sequencing of advanced gynaecological malignancies in National University Cancer Institute, Singapore (NCIS). We retained cases with successful sequencing on FoundationOne CDx and an evaluable response to first line treatment. Patients who had undergone upfront cytoreductive surgery with no remaining evaluable disease post-operatively were excluded. Regimens in retained cases included cis/carbo-platin (combined as for analysis purposes) and paclitaxel.

Both IMAC-CRC and IMAC-OV included cases utilising the FoundationOne CDx testing platform, giving mutational information of 324 genes of interest. We included reported pathogenic alterations and variants of uncertain significance.

#### Cell Lines

The raw data was obtained from the CCLE (Barretina 2012) DepMap portal (https://depmap.org/portal/download/all/?releasename=DepMap+Public+21Q3). The drug response for these cell lines was obtained from the GDSC portal (https://www.cancerrxgene.org/downloads/bulk_download). We retained the cell lines which have a corresponding drug response measured in terms of AUDRC.

For training with datasets listed above (TCGA, IMAC-CRC and IMAC-OV) we filter cell lines with responses to drugs retained in the the dataset in question.

- For training with TCGA, we filter cell lines with responses to the drugs Cisplatin, Paclitaxel, 5-Fluorouracil, Gemcitabine, Docetaxel and Cyclophosphamide. This set of cell lines and TCGA patient samples is labelled CCLE-TCGA dataset.
- For training with IMAC-CRC, we filter cell lines with responses to the drugs 5-Fluorouracil, Irinotecan, Cetuximab and Oxaliplatin. We call this set of cell lines and IMAC-CRC samples the CCLE-CRC dataset.
- For training with IMAC-OV samples, we filter cell lines with responses to the drugs Cisplatin, Paclitaxel, Gemcitabine and Doxorubicin. This set of cell lines and IMAC-OV samples is labelled the CCLE-OV dataset.

The CCLE-TCGA dataset consists of 689 cell lines and 470 TCGA patients, the CCLE-CRC dataset contains 689 cell lines and 82 colorectal cancer patients, while the CCLE-OV dataset contains 677 cell lines and 105 ovarian cancer patients.

##### Mutations

We retain only the mutations that are annotated as “damaging” and “other non-conserving”.

##### Gene Expression

The gene expression data from the CCLE DepMap portal was used directly.

### Experiment Settings

After processing described above, we have three datasets consisting of a combination of cell lines and patient samples: (1) CCLE-TCGA, (2) CCLE-CRC and (3) CCLE-OV. The 3 datasets (i.e. (patient, drug) pairs) were divided into 80-20 train-test splits, with 3 different random states to generate 3 splits (Supplementary Tables 5-15).

#### Evaluation Metrics

Since we modelled the task of drug response prediction in patients as a classification problem, we evaluated the performance of the model in terms of Area Under the Receiver Operating Characteristic curve (AUROC) and the Area Under the Precision Recall Curve (AUPRC). These metrics were calculated on the held-out 20% test splits on all 3 splits for CCLE-TCGA, CCLE-OV and CCLE-CRC datasets. The overall AUROC and AUPRC were calculated for each split by considering the predictions and ground truth labels for all the drugs together. The final results were reported per drug on each dataset, for all those drugs with at least 80 RECIST responses for CCLE-OV and CCLE-TCGA datasets, and at least 10 RECIST responses for CCLE-CRC dataset. The baseline AUPRC is calculated as the fraction of positive labelled test (patient, drug) pairs with respect to all test (patient, drug) pairs (Saito and Rehmsmeier).

#### Features

We encode the mutation, gene expression and copy number variation data, for all datasets, into vectors of different dimensions (Supplementary Table 16). These vectors can be binary (CNV, mutations) or real valued(annotated mutations, gene expression). These vectors also differ in the dimensionality based on the input genes used and the processing done.

#### Experiments

##### Clinical NGS data is sufficient for DRP model performance

CODE-AE was trained on the train split of the CCLE-TCGA dataset (comprising patients and cell lines) and evaluated on the patients in the test split. 3 subsets of genes were considered:

i. only FoundationOne cNGS panel genes (324 genes)
ii. whole exome sequencing (WES) panel of 19536 genes and
iii. 285 genes that are common across FoundationOne, TruSight Oncology 500 and Tempus xF+ panels.

This was repeated across all 3 random train-test splits of the CCLE-TCGA dataset. Details of the input features in each subset are provided in Supplementary Table 16.

For WES, the feature space dimension was first reduced by using an autoencoder (AE) to project down to 324 dimensions before running CODE-AE. The AE had an encoder-decoder architecture with one bottleneck layer to project from 19536 to 324 dimensions. It was trained to minimise the Mean Squared Error (MSE) loss between input and reconstructed matrices, over 2000 epochs with a learning rate of 1e-4 and convergence threshold of 1e-5. This allowed us to train in our computing environment that had limited memory. For evaluation, the overall AUROC and AUPRC were calculated for each train-test split, by combining the predictions for each (patient, drug) pair in the test split. During evaluation only (patient, drug) pairs with 5-Fluorouracil, Cisplatin and Paclitaxel as the drug were considered. These drugs had more than 80 (patient, drug) pairs in the TCGA dataset. To calculate the overall AUROC and AUPRC, the predicted responses from CODE-AE for (patient, drug) pairs for all three drugs, 5-Fluorouracil, Cisplatin and Paclitaxel, and their ground truth RECIST labels, were considered together. To test if the performance was significantly different across the 3 gene subsets, we conducted an ANOVA test across the overall AUROC and AUPRC for the 3 test splits. This resulted in a p-value of 0.8367 for overall AUROC and 0.78 for overall AUPRC. As such, we could not reject the null hypothesis that all 3 subsets had a similar performance across 3 test splits.

Further, we combined all (patient, drug) pairs in the test splits of the 3 train-test splits, with respect to the predicted responses from CODE-AE and the ground truth RECIST labels. For (patient, drug) test pairs present in more than one test split, we took the mean predicted response across the test splits. This aggregation allowed us to combine the test split (patient, drug) pairs across the 3 test splits. In total the aggregated test set had 203 samples (patient, drug pairs on which the model predicts), with 90, 82 and 90 pairs across the three splits. To consider the differences across cancer types, we considered cancer types with more than 20 (patient, drug) test pairs. We repeated the above comparison across all 3 subsets of genes, using Velodrome as well(Supplementary Fig. 2).

##### DruID: predicting chemotherapy drug response with cNGS data

All the existing baseline models (CODE-AE, TCRP, TUGDA, Velodrome) were trained on the cell lines and patients in the train splits of the CCLE-TCGA dataset, and evaluated on the patients in the corresponding test splits. Only 324 genes from the FoundationOne panel were considered for all the experiments. This was repeated across all 3 train-test splits of the CCLE-TCGA dataset. For the baseline methods, the inputs were binary mutation vectors (i.e., without variant annotations). For DruID, initially a model was trained using CCLE-OV data (including all IMAC-GO patients) (annotation, unsupervised domain adaptation and multi-task learning). Then another DruID model was instantiated with these learnt weights for each of the 3 drugs - 5-Fluorouracil, Cisplatin and Paclitaxel for each train split. Each of these drug-split-specific models was trained using the CCLE-TCGA train split consisting of (patient,drug) train pairs where the drug matched the drug in the drug-split-specific model.

For evaluation, the overall AUROC and AUPRC were calculated for each train-test split, by combining the predictions for each (patient, drug) pair in the test split. To calculate the overall AUROC and AUPRC, the predicted responses from all the baseline methods for (patient, drug) pairs with 5-Fluorouracil, Cisplatin and Paclitaxel, along with their ground truth RECIST labels, were considered together. We checked the significance of overall AUROC and AUPRC across the 3 test splits for DruID and Velodrome (second-best performing model), using a t-test. We obtained a p-value of 0.004 on AUROC and 0.037 on AUPRC, indicating a significant difference between the performance of DruID and the second best-performing model.

Further, for each method, we combined all (patient, drug) pairs in the 3 test splits as described earlier.The aggregated test set had 203 samples (patient-drug pairs), with 90, 82 and 90 pairs across the three splits. This aggregated test set had 88 patients treated with Cisplatin, 58 patients treated with Paclitaxel and 57 patients treated with 5-Fluorouracil.

We also conducted an ablation study by successively removing components of the DruID architecture(Supplementary Fig. 3). In the first ablation, we removed the variant annotation step. In the second ablation, we also removed the zero inflated loss terms and the zero inflated layer in the unsupervised domain adaptation step.

##### Copy number variation (CNV) information or gene expression data does not improve DruID performance

DruID was trained with different input data types, in this experiment. We used the cell lines and patients in the train splits of the CCLE-TCGA dataset for training and evaluated it on the patients in the test splits.

We compared the performance when using only CNV data (this was one hot encoded to indicate loss, no change and amplification), only variant annotated mutation data and a combination of the two. For combining binary CNV and real valued variant annotated mutation data, the UDA step involved the use of 2 separate ZI VAEs (ZINB for CNV and ZINorm for variant annotated mutations) per domain. The representations from both ZI VAEs were concatenated and used as the representation in the further layers of the architecture. For evaluation, the overall AUROC and AUPRC were calculated for each train-test split, by combining the predictions for each (patient, drug) pair in the test split. To check the significance of the performance using annotated mutations, we ran a t-test between AUROC and AUPRC from DruID trained using only variant annotated mutations and DruID trained using a combination of copy number variation and variant annotated mutations (second best performing model), across the 3 test splits. This yielded a p-value of 0.003 for AUROC and 0.013 for AUPRC, which indicated significant difference in performance of annotated mutations over copy number variation.

Further, for each method, we combined all (patient, drug) pairs in the 3 test splits to obtain an aggregated test split, as described earlier, to obtain the AUROC and AUPRC curves (Fig. 3 (a)). We also compared the performance of these 3 input data types, across various drugs. For each input type, the mean AUROC and AUPRC for each drug across all 3 test splits were calculated, while plotting Fig. 3 (b). The significance was tested by comparing the AUROC and AUPRC across the 3 test splits for each drug, between annotated mutations and a combination of annotated mutations and copy number variation using a t-test. Annotated mutations were significantly better with respect to AUPRC and AUROC for 5-Fluorouracil (*p* = 0.004), and for AUROC for Paclitaxel (*p* = 0.009).

Further, we compared the performance when using only gene expression data, only variant annotated mutation data and a combination of the two. Both data types were real valued and involved the use of ZINorm VAEs. Since some TCGA samples did not have gene expression data available, we dropped these from the test splits while comparing the performance across the various input types. For evaluation, the overall AUROC and AUPRC were calculated for each train-test split, by combining the predictions for each (patient, drug) pair in the test split. To check the significance of the performance using annotated mutations, we ran a t-test between AUROC and AUPRC from DruID trained using only variant annotated mutations and DruID trained using only gene expression (second best performing model), across the 3 test splits. This yielded a p-value of 0.007 for AUROC and 0.04 for AUPRC, which indicated significant difference in performance of annotated mutations over gene expression. Similar to CNV, we combined all (patient, drug) pairs in the test splits of the 3 train-test splits, with respect to the predicted responses from the method and the ground truth RECIST labels, to obtain the AUROC and AUPRC curves in Fig. 3(c).

We also compared the performance of these 3 input data types, across various drugs. The significance was tested by comparing the AUROC and AUPRC across the 3 test splits for each drug, between annotated mutations and gene expression using a t-test. Annotated mutations were significantly better with respect to AUPRC (*p* = 0.021) and AUROC for Cisplatin (*p* = 0.028). Details of the input feature vectors are in Supplementary Table 16.

##### Validating DruID on clinical datasets

We evaluated DruID on two clinical datasets - CCLE-CRC and CCLE-OV, as described in the Results section.

DruID was trained using the train splits of each dataset and evaluated on the corresponding test splits. We combined all (patient, drug) pairs in the 3 test splits (34, 33, 35 patient, drug pairs in each test split of CCLE-CRC and 32, 64, 32 patient, drug pairs in each test split of CCLE-OV dataset) as described earlier. The aggregated CRC test set had 38 patients treated with 5-Fluorouracil, 23 patients treated with Oxaliplatin and 15 patients treated with Irinotecan, resulting in a total of 76 test (patient, drug) pairs. The aggregated OV test set had 55 patients treated with Cisplatin/Carboplatin, 53 patients treated with Paclitaxel, resulting in a total of 108 test (patient, drug) pairs. The mean predicted probability of response (Figure 4(d)), across responders and non-responders to each drug, was calculated by passing the prediction from DruID through a sigmoid function.

Oncoplots for predicted non-responders and responders to 5FU and Cisplatin/carboplatin, across 3 train-test splits on patients in CCLE-CRC and CCLE-OV, were generated using the maftools R package (Mayakonda 2018) (Fig. 4(e, f)). Predicted responders were those with predicted response in the top 20th percentile of predicted responses to the drug and predicted non-responders were those with predicted response in the bottom 20th percentile of predicted responses to the drug.

## Acknowledgements and Funding

David SP Tan is supported by the National Medical Research Council, Singapore under its NMRC Clinician Scientist Award (MOH-001006) and has received charitable research funding from the Pangestu Family Foundation Gynaecological Cancer Research Fund. The ongoing IMAC study is supported by National Research Foundation, Singapore and National Medical Research Council, Singapore under its NMRC Centre Grant Programme (NMRC/CG/M005/2017_NCIS).

Vaibhav Rajan acknowledges support from ‘AI Singapore 100 Experiments’ Grant No. AISG-100E-2023-116 (PI: Vaibhav Rajan).

Aishwarya Jayagopal is supported by the National University of Singapore Research Scholarship.

## Data Availability

TCGA and CCLE datasets are publicly available as referenced in main text. The remaining datasets generated during and/or analysed during the current study are available from the corresponding author on reasonable request.

## Code Availability

Code is available at https://github.com/CDAL-SOC/druid_paper.git - branch ‘v2.0’

## Figure Creation

Images in this paper were created using FlatIcon and Freepik.

## Supplementary Material

### Background on other ML approaches

Prior literature on drug response prediction has largely focused on cancer cell lines(Adam 2020, Chen 2021, Firoozbakht 2021). The availability of transcriptomic data in the form of gene expression, mutations, copy number variations in cell lines has resulted in a wide variety of machine learning models for drug response prediction. These methods range from linear regression and ensemble models to graph neural networks. However, DRP models trained on cell lines alone often translate poorly to patients (Mourragui 2019, Mourragui 2021, Sharifi-Noghabi 2020). This is partly due to inherent biological differences, meaning cell lines do not accurately represent patient tumours. Cell lines are essentially a subpopulation of the primary tumour and do not exhibit heterogeneity seen *in vivo*. The absence of the tumour microenvironment and interactions with the host of stromal cells present in patients is also key (Mourragui 2019, Huo 2020). In addition, technical differences in response measurement in cell lines versus in patients, and differences in drug dosing between cell lines and patients will affect interpretation of results by a DRP model.

While omics data is increasingly available for many cancer patients (TCGA 2013, Cerami 2012), drug response data for these patients remains scarce and limited to standard of care therapies only. To address such challenges, transfer learning approaches including domain adaptation have been developed to train DRP models from both cell lines and patients (He 2022, Sharifi 2021, Ma 2021). Transfer learning approaches are useful when there are limited samples available in the domain of interest (target domain), but a related domain (source domain) has a large number of labelled samples. (Pan 2009) have broadly grouped various transfer learning approaches based on whether the source and target domains are labelled or not. When both source and target domains are unlabeled, it is called unsupervised transfer learning. Methods which learn a shared representation space as part of pretraining, like CODE-AE (He 2022), fall into this category. If the source domain has labelled samples but the target domain is unlabeled, it is called transductive transfer learning. Methods like TUGDA, PACE, Velodrome (Peres 2021, Anastopoulos 2021, Sharifi 2021) fall into this category. If both domains have labelled samples, it is called inductive transfer learning. Methods like AITL (Sharifi 2020, Prasse 2022), TCRP (Ma 2021), molecular pathway based model (Tang 2022) use this approach where they utilise the limited number of labelled target domain samples as well.

In all of these methods, they focus on one of two aspects that differentiate patients from cell lines - (1) distributional differences in omic profiles owing to differences in biological environment - termed “input space discrepancy”and (2) differences in the way drug response is measured - termed “output space discrepancy” (Sharifi 2020). Input space discrepancy is handled by finding a shared embedding space that is common to both cell lines and patients. To deal with the output space discrepancy, one way is to have separate drug response prediction networks for cell lines and patients. Doing so allows both networks to learn nuances specific to each domain, in the output space. Except AITL and Velodrome, all other methods address only one of these two discrepancies. Models like (Prasse 2022), (Tang 2022), CODE-AE (He 2022), PACE (Anastopoulos 2021), PRECISE (Mourragui 2019), TRANSACT (Mourragui 2021), TUGDA (Peres 2021) and TCRP (Ma 2021) handle input space discrepancy but the same drug response prediction network is used by both the domains. The discrepancy in the output space is handled either by discretizing the cell line response based on empirically determined thresholds or by evaluating the predictions using correlation metrics. Models like Velodrome and AITL use a shared space for the domains to address input space discrepancy and use two separate prediction networks to handle discrepancies in the output space. In both cases, they train one model for each drug. When networks are trained in this manner, it becomes difficult to predict the response for a new/unseen drug since there is no trained network to perform inference with. This proves to be a challenge in the problem of drug repurposing.

State-of-the-art transfer learning methods, which evaluated their models on patient data, have largely restricted their analysis to gene expression data (Sharifi 2021, Peres 2021). For example, CODE-AE(He 2022) used a set of 1426 genes (selected based on percentage of unique gene expression values) and Velodrome(Sharifi 2021) used a set of 2128 genes (selected based on network propagation over a protein-protein interaction network). The genes selected in these methods are not captured based on their presence in cNGS panels; nor are the number of chosen genes comparable across cNGS and these methods. Moreover, unlabeled patient samples remain unused resulting in inefficient use of available data. Requiring transcriptomic input data represents a challenge in bringing these methods to mainstream patient care and it remains unknown if such tools can accurately predict response from the limited number of recurrently altered cancer genes that are included in cNGS panels such as FoundationOne CDx (324 genes), Tempus (523 genes), and TruSight Oncology 500 (523 genes). To the best of our knowledge, no prior transfer learning methods have been evaluated on such a restricted subset of genes. Moreover, methods which have used mutations as inputs, have not considered the variant level information captured in cNGS reports; instead they treat all alterations as equal, resulting in loss of granularity and potential reduction in predictive accuracy. A comparison of existing DRP methods is available in Table 2.

### Supplementary Results

**Supplementary Figure 1:**
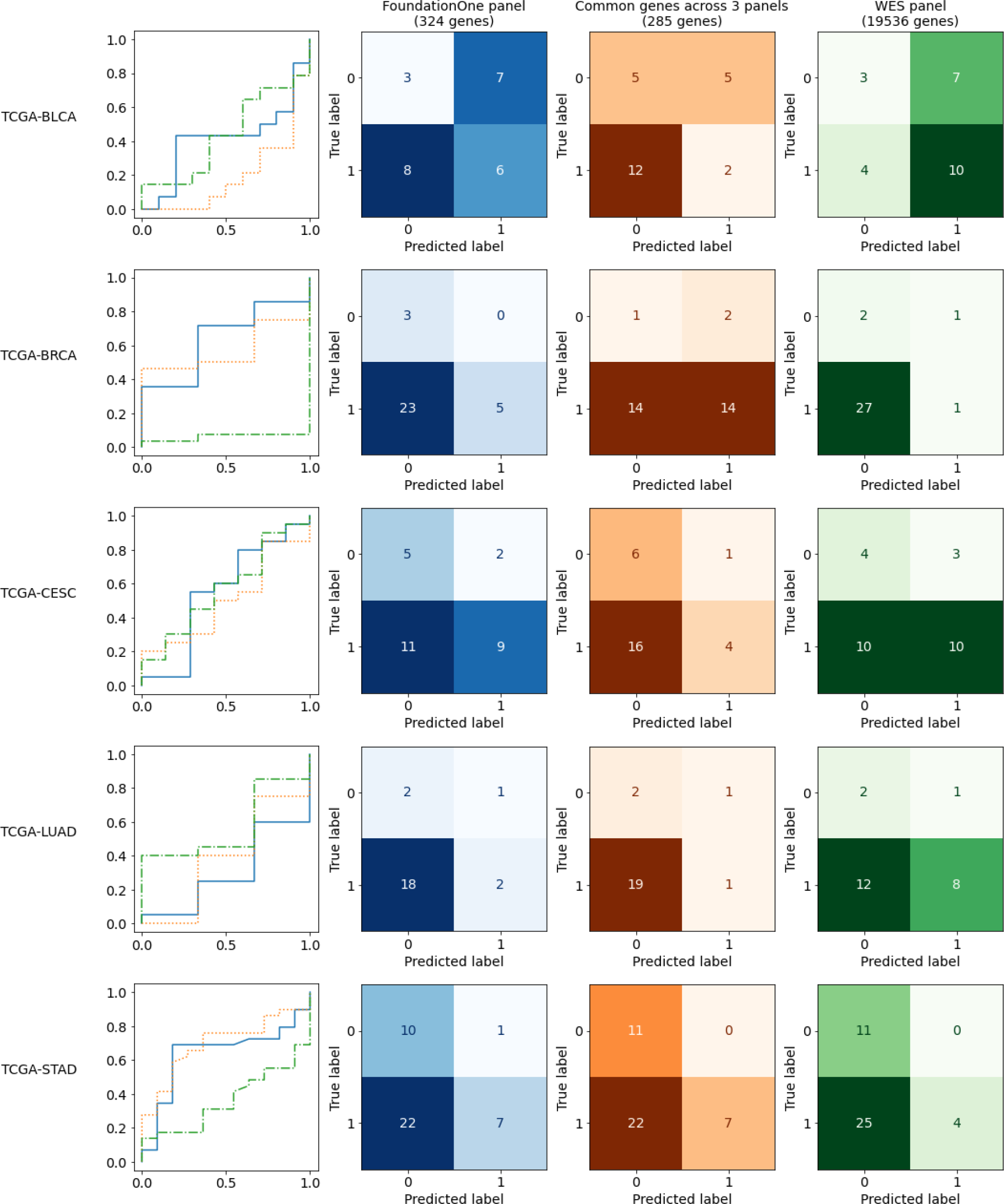
Comparison of CODE-AE performance across 3 cNGS panels distinguished by cancer type (BLCA: Bladder Urothelial Carcinoma, BRCA: Breast invasive carcinoma, CESC: Cervical squamous cell carcinoma and endocervical adenocarcinoma, LUAD: Lung adenocarcinoma, STAD: Stomach adenocarcinoma, UCEC: Uterine Corpus Endometrial Carcinoma). Only cancer types with more than 20 (patient, drug) test pairs are considered here. In most cancer types, the confusion matrices look similar across all 3 cNGS panels suggesting a similar predictive performance of cNGS panels compared to WES.

**Supplementary Table 1:** CODE-AE performance across various subsets of genes.

|  | Foundation One<br>(324 genes) | Common genes<br>across 3 panels<br>(285 genes) | WES panel<br>(19,536 genes) |
| --- | --- | --- | --- |
| Sensitivity/Recall | 0.272 | 0.291 | 0.311 |
| Specificity | 0.673 | 0.692 | 0.692 |
| Precision | 0.707 | 0.733 | 0.746 |

Sensitivity, Specificity and Precision values, from CODE-AE, corresponding to confusion matrices in Fig.1b.

**Supplementary Figure 2 (a).**
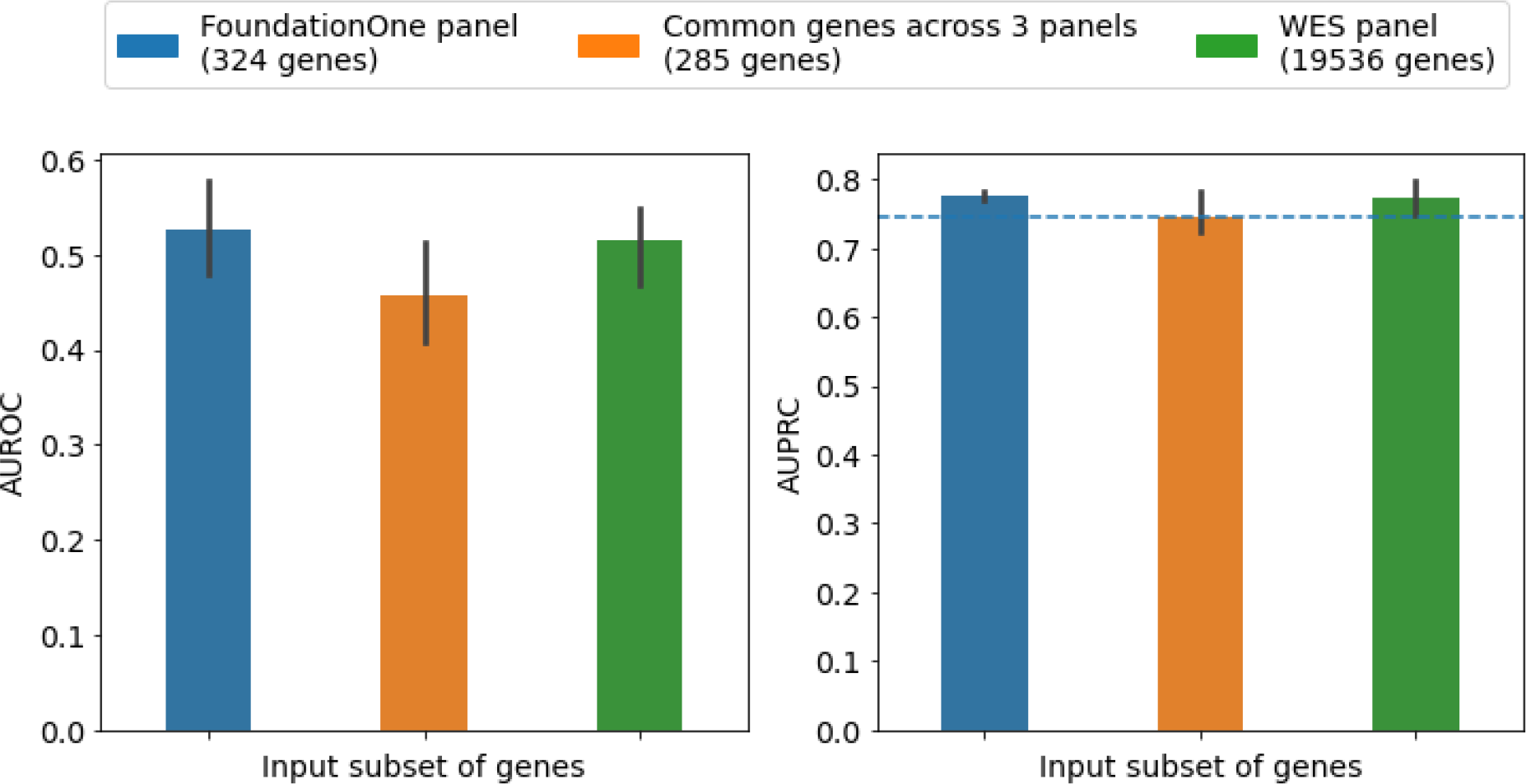
Comparison of AUROC and AUPRC scores of response prediction for different input subsets of genes. Performance is measured on 3 randomly chosen test splits, using TCGA data. Velodrome is used to predict response. Results show that performance is not significantly different (p-value associated with AUROC comparison: 0.259, p-value associated with AUPRC comparison: 0.281) across the 3 subsets of genes, suggesting that predictive value of a subset of genes used in cNGS panels is similar to that of all genes from WES. Baseline value for AUPRC: 0.7438

**Supplementary Figure 2 (b).**
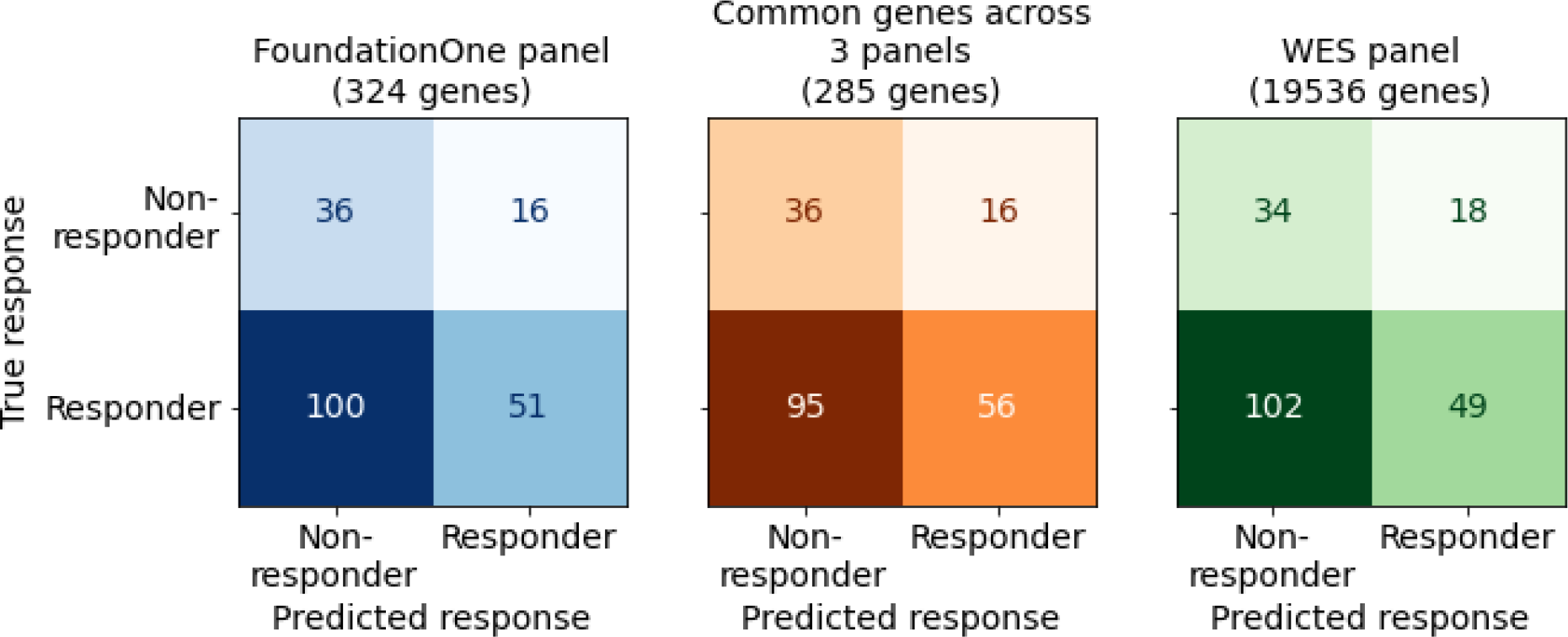
Confusion matrices for different input subsets of genes on 203 samples from TCGA; predictions obtained using the method Velodrome. Predicted values were converted to binary responses using FPR and TPR thresholds of 0.3 each. Colour indicates the input subset, shade indicates magnitude of the values. All 3 subsets of genes yield similar distributions in the confusion matrices.

**Supplementary Table 2:** Velodrome performance across various subsets of genes.

|  | Foundation One<br>(324 genes) | Common genes<br>across 3 panels<br>(285 genes) | WES panel<br>(19,536 genes) |
| --- | --- | --- | --- |
| Sensitivity/Recall | 0.3377 | 0.3709 | 0.3245 |
| Specificity | 0.6923 | 0.6923 | 0.6538 |
| Precision | 0.7612 | 0.7778 | 0.7313 |

Sensitivity, Specificity and Precision values, from Velodrome, corresponding to confusion matrices in Figure Supplementary Fig. 2 (b).

**Supplementary Figure 3.**
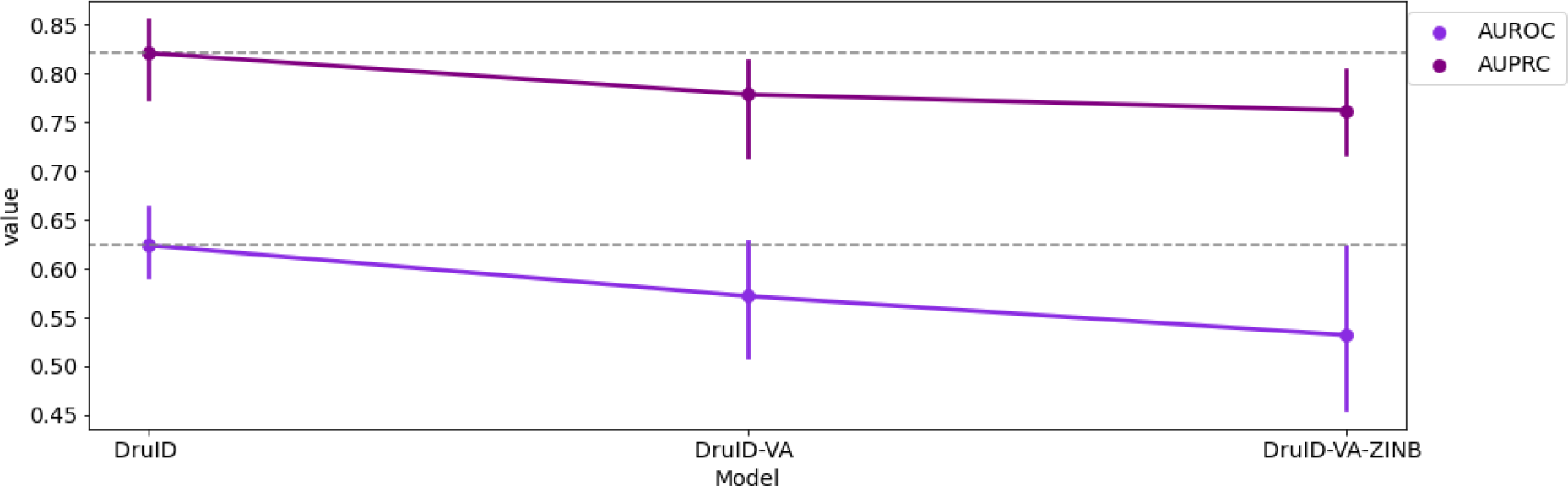
Ablation study with DruID. Performance (AUPRC, above and AUROC, below) after incremental component-wise removal from DruID of variant annotation (DruID-VA) and zero inflated loss (DruID-VA-ZINB). Removal of each component reduces DruID’s performance, thus showing the importance of each component.

Supplementary Fig. 3 shows the importance of two of our modelling strategies through an ablation study. From the complete DruID method, we first remove stage I to obtain DruID-VA (“DruID minus VA”). In DruID-VA, variant annotations are not used; instead 324-dimensional binary representations of cell lines and patients are used with each binary value indicating presence/absence of mutation(s) in the gene. We observe that DruID-VA achieves AUROC, AUPRC of 0.5715 and 0.7783 respectively, which is lower than that of DruID (0.6236 and 0.8206 respectively). Next, we change our VAE model, to not use zero-inflated distributions to model sparse inputs. This further reduces the performance to AUROC of 0.5316 and AUPRC of 0.762.

### Data Processing

**Supplementary Table 3:** Mapping ClinVar annotations.

| Updated category | ClinVar generated annotations |
| --- | --- |
| Pathogenic | Pathogenic, Pathogenic\|drug_response\|other, Pathogenic/Likely_pathogenic, Likely_pathogenic, Pathogenic/Likely_pathogenic\|other, drug_response, Likely_pathogenic\|other, Pathogenic\|risk_factor, Pathogenic/Likely_pathogenic\|drug_response, Likely_risk_allele, risk_factor |
| Benign | Likely_benign, Benign/Likely_benign, Benign |
| Variants of Unknown Significance | ., Uncertain_significance, Conflicting_interpretations_of_pathogenicity, not_provided, Conflicting_interpretations_of_pathogenicity\|other, Uncertain_significance\|drug_response, other |

Mapping of clinical significance categories obtained from ClinVar to 3 broad annotation categories - pathogenic, benign and variants of unknown significance.

**Supplementary Table 4:**
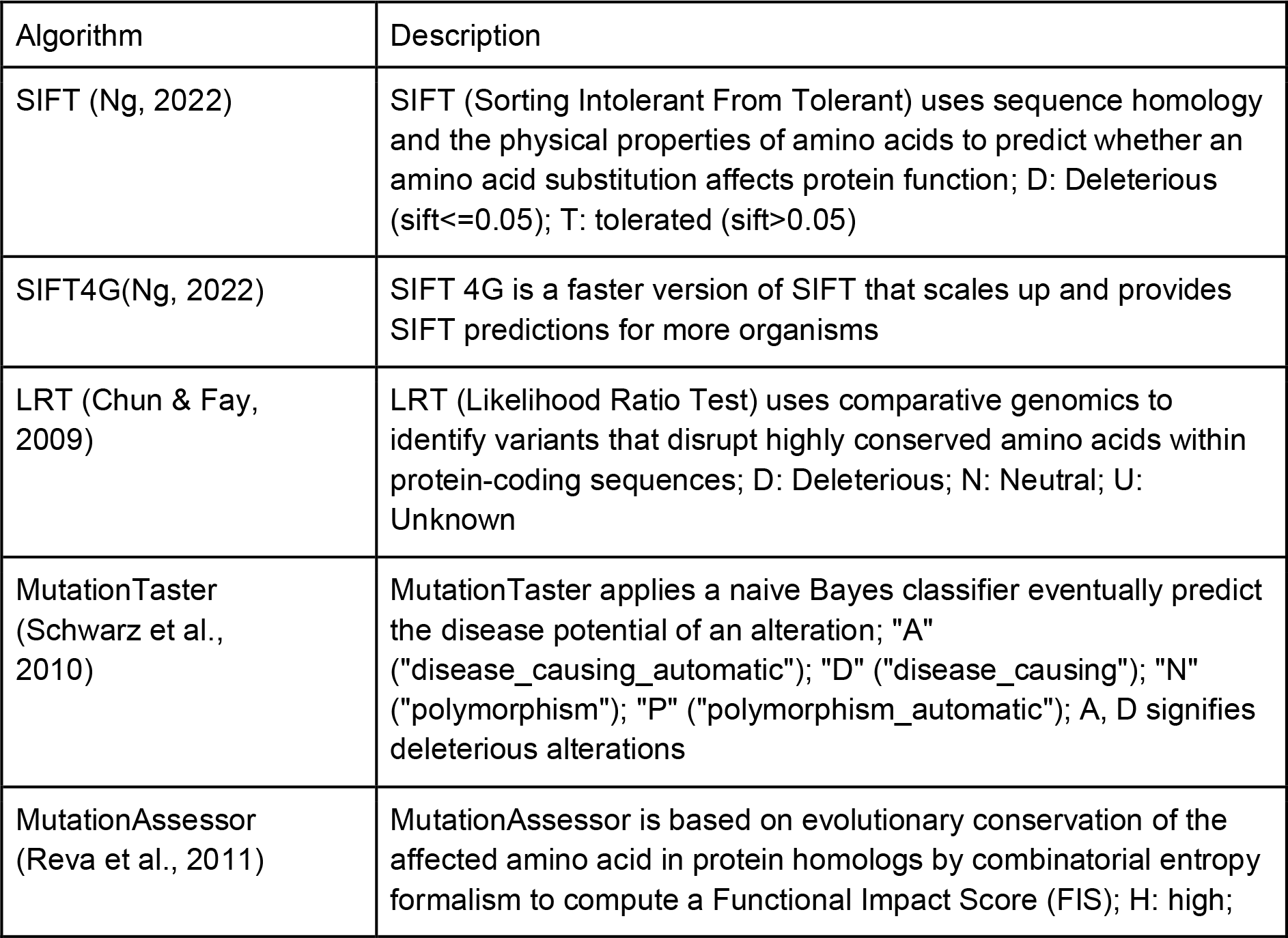
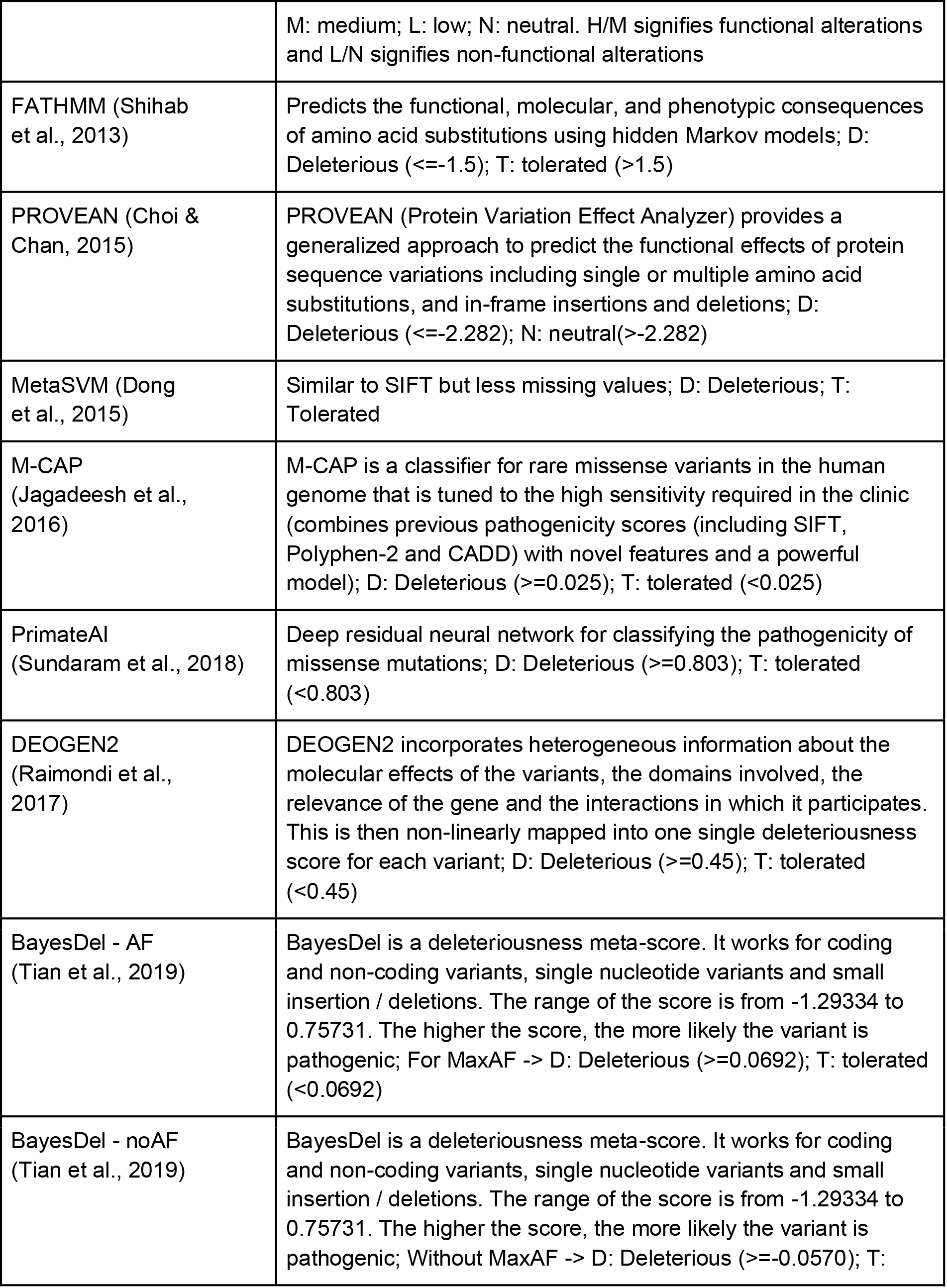
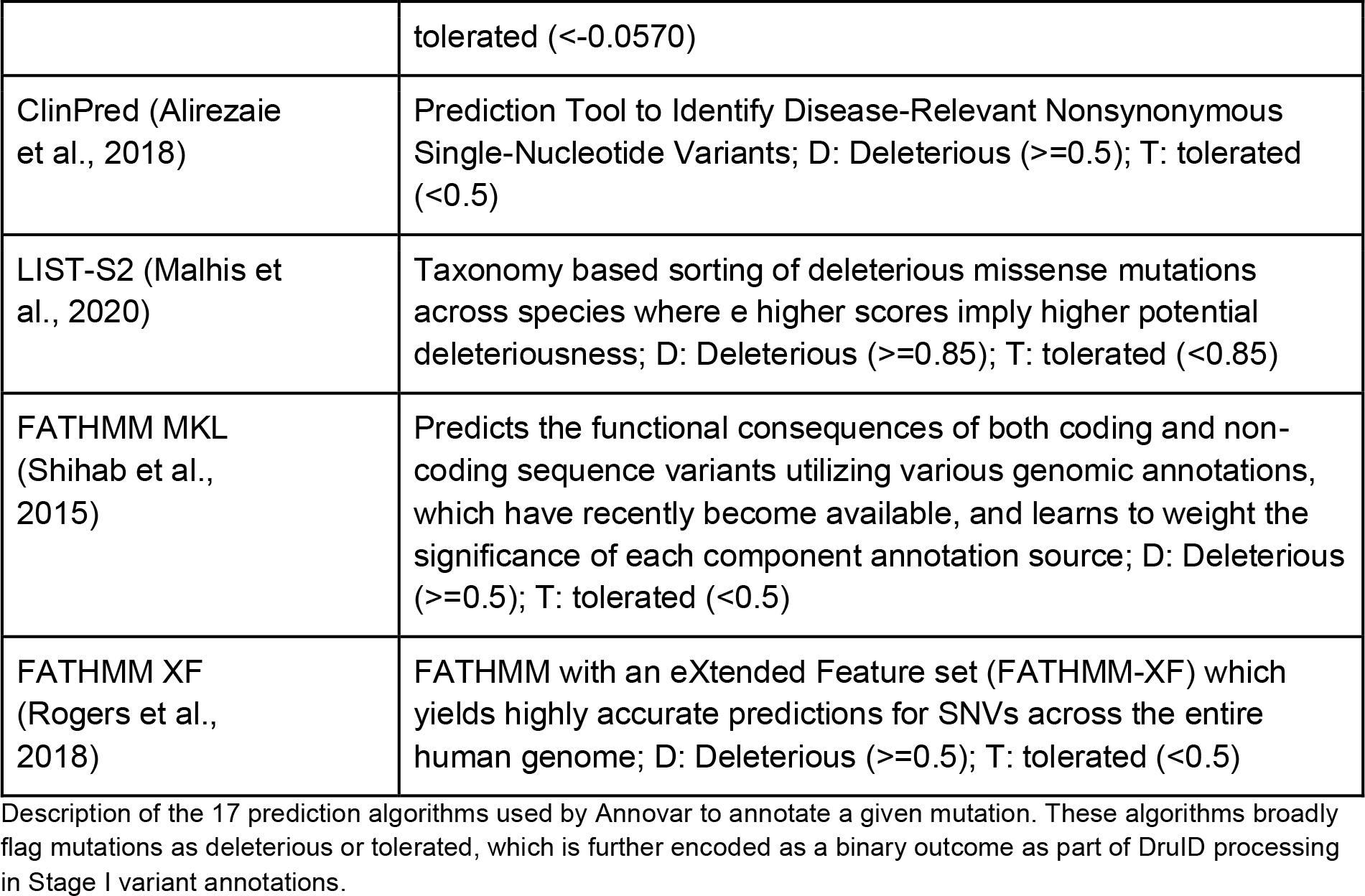
Description of AnnoVar annotations.

### Data Statistics

**Supplementary Table 5.**

| Drug Name | Train split<br>TCGA | Test split<br>TCGA | Train split<br>CCLE | Test split<br>CCLE |
| --- | --- | --- | --- | --- |
| CISPLATIN | 167 | 39 | 425 | 112 |
| PACLITAXEL | 87 | 26 | 542 | 134 |
| 5-FLUOROURA<br>CIL | 100 | 25 | 468 | 121 |
| OVERALL | 354 | 90 | 1435 | 367 |

Number of (patient, drug) pairs in the train and test splits of CCLE-TCGA dataset split 0. Each cell indicates the number of patients/samples who were treated with the corresponding drug, which are further divided up across the train and test splits.

**Supplementary Table 6:** Overall - cancer type distribution:

| Cancer type | Train split TCGA | Test split TCGA |
| --- | --- | --- |
| BLCA | 48 | 8 |
| BRCA | 56 | 21 |
| CESC | 49 | 13 |
| COAD | 27 | 7 |
| HNSC | 46 | 7 |
| LUAD | 39 | 12 |
| LUSC | 22 | 3 |
| SKCM | 10 | 1 |
| STAD | 59 | 16 |
| UCEC | 27 | 7 |

Supplementary Table 7a: Number of (patient, drug) pairs in the train and test splits of TCGA dataset split 0. Each cell indicates the number of patients belonging to the specific cancer type indicated by the row, which are further divided up across the train and test splits.

#### Drug specific - cancer type distribution (TCGA)

**Supplementary Table 7:** Number of (patient, drug) pairs in the train and test splits of TCGA dataset split 0. Each cell indicates the number of patients belonging to the specific cancer type indicated by the row, which are further divided up across the train and test splits. These are also categorised based on the drug administered in each case.

| Drug | Cancer type | Train split TCGA | Test split TCGA |
| --- | --- | --- | --- |
| CISPLATIN | BLCA | 32 | 4 |
|  | CESC | 39 | 11 |
|  | HNSC | 32 | 6 |
|  | LUAD | 27 | 10 |
|  | LUSC | 14 | 1 |
|  | SKCM | 7 | 0 |
|  | STAD | 12 | 7 |
|  | UCEC | 4 | 0 |
| PACLITAXEL | BLCA | 8 | 0 |
|  | BRCA | 19 | 12 |
|  | CESC | 6 | 2 |
|  | HNSC | 12 | 1 |
|  | LUAD | 11 | 2 |
|  | LUSC | 2 | 1 |
|  | SKCM | 1 | 1 |
|  | STAD | 3 | 0 |
|  | UCEC | 25 | 7 |
| 5-FLUOROURACIL | BLCA | 1 | 0 |
|  | BRCA | 16 | 5 |
|  | CESC | 4 | 0 |
|  | COAD | 27 | 7 |
|  | HNSC | 1 | 0 |
|  | STAD | 51 | 13 |

#### Split 0 (CCLE/NUH CRC)

**Supplementary Table 8:**
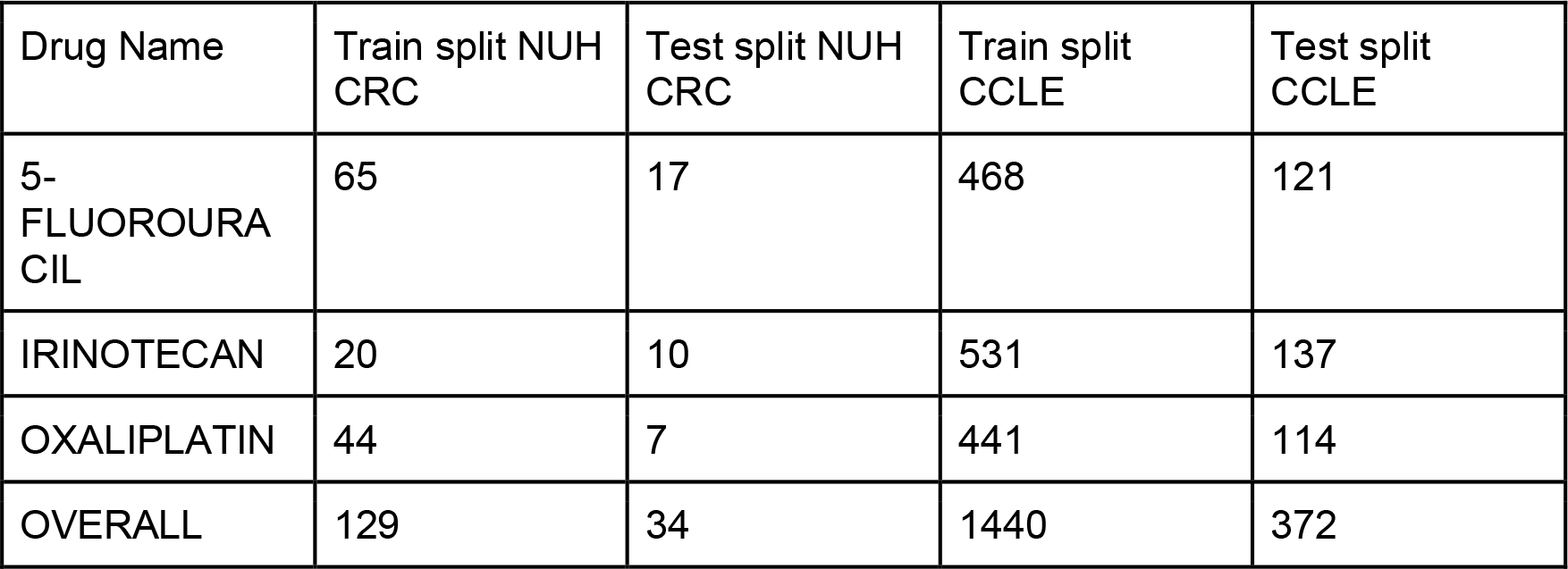
Number of (patient, drug) pairs in the train and test splits of CCLE-NUH CRC dataset split 0. Each cell indicates the number of patients/samples who were treated with the corresponding drug, which are further divided up across the train and test splits.

#### Split 0 (CCLE/NUH Ovarian)

**Supplementary Table 9:** Number of (patient, drug) pairs in the train and test splits of CCLE-NUH ovarian dataset split 0. Each cell indicates the number of patients/samples who were treated with the corresponding drug, which are further divided up across the train and test splits.

| Drug Name | Train split NUH Ovarian | Test split NUH Ovarian | Train split CCLE | Test split CCLE |
| --- | --- | --- | --- | --- |
| CISPLATIN | 88 | 17 | 425 | 112 |
| PACLITAXEL | 87 | 15 | 540 | 136 |
| GEMCITABINE |  |  | 425 | 112 |
| DOXORUBICIN |  |  | 447 | 110 |
| OVERALL | 175 | 32 | 1837 | 470 |

#### Split 1 (CCLE/TCGA)

**Supplementary Table 10:** Number of (patient, drug) pairs in the train and test splits of CCLE-TCGA dataset split 1. Each cell indicates the number of patients/samples who were treated with the corresponding drug, which are further divided up across the train and test splits.

| Drug Name | Train split TCGA | Test split TCGA | Train split CCLE | Test split CCLE |
| --- | --- | --- | --- | --- |
| CISPLATIN | 168 | 38 | 426 | 111 |
| PACLITAXEL | 87 | 26 | 538 | 138 |
| 5-FLUOROURACIL | 107 | 18 | 472 | 117 |
| OVERALL | 362 | 82 | 1436 | 366 |

#### Split 1 (CCLE/NUH CRC)

**Supplementary Table 11:** Number of (patient, drug) pairs in the train and test splits of CCLE-NUH CRC dataset split 1. Each cell indicates the number of patients/samples who were treated with the corresponding drug, which are further divided up across the train and test splits.

| Drug Name | Train split NUH CRC | Test split NUH CRC | Train split CCLE | Test split CCLE |
| --- | --- | --- | --- | --- |
| 5-FLUOROURACIL | 65 | 17 | 468 | 121 |
| IRINOTECAN | 22 | 8 | 532 | 136 |
| OXALIPLATIN | 43 | 8 | 440 | 115 |
| OVERALL | 130 | 33 | 1440 | 372 |

#### Split 1 (CCLE/NUH Ovarian)

**Supplementary Table 12:** Number of (patient, drug) pairs in the train and test splits of CCLE-NUH Ovarian dataset split 1. Each cell indicates the number of patients/samples who were treated with the corresponding drug, which are further divided up across the train and test splits.

| Drug Name | Train split NUH Ovarian | Test split NUH Ovarian | Train split CCLE | Test split CCLE |
| --- | --- | --- | --- | --- |
| CISPLATIN | 73 | 32 | 430 | 107 |
| PACLITAXEL | 70 | 32 | 540 | 136 |
| GEMCITABINE | 1 |  | 430 | 107 |
| DOXORUBICIN |  |  | 449 | 108 |
| OVERALL | 144 | 64 | 1849 | 458 |

#### Split 2 (CCLE/TCGA)

**Supplementary Table 13:** Number of (patient, drug) pairs in the train and test splits of CCLE-TCGA dataset split 2. Each cell indicates the number of patients/samples who were treated with the corresponding drug, which are further divided up across the train and test splits.

| Drug Name | Train split<br>TCGA | Test split<br>TCGA | Train split<br>CCLE | Test split<br>CCLE |
| --- | --- | --- | --- | --- |
| CISPLATIN | 167 | 39 | 431 | 106 |
| PACLITAXEL | 86 | 27 | 541 | 135 |
| 5-FLUOROURA<br>CIL | 101 | 24 | 473 | 116 |
| OVERALL | 354 | 90 | 1445 | 357 |

#### Split 2 (CCLE/NUH CRC)

**Supplementary Table 14:** Number of (patient, drug) pairs in the train and test splits of CCLE-NUH CRC dataset split 2. Each cell indicates the number of patients/samples who were treated with the corresponding drug, which are further divided up across the train and test splits.

| Drug Name | Train split NUH<br>CRC | Test split NUH<br>CRC | Train split<br>CCLE | Test split<br>CCLE |
| --- | --- | --- | --- | --- |
| 5-FLUOROURA<br>CIL | 65 | 17 | 473 | 116 |
| IRINOTECAN | 24 | 6 | 534 | 134 |
| OXALIPLATIN | 39 | 12 | 451 | 104 |
| OVERALL | 128 | 35 | 1458 | 354 |

#### Split 2 (CCLE/NUH Ovarian)

**Supplementary Table 15:**
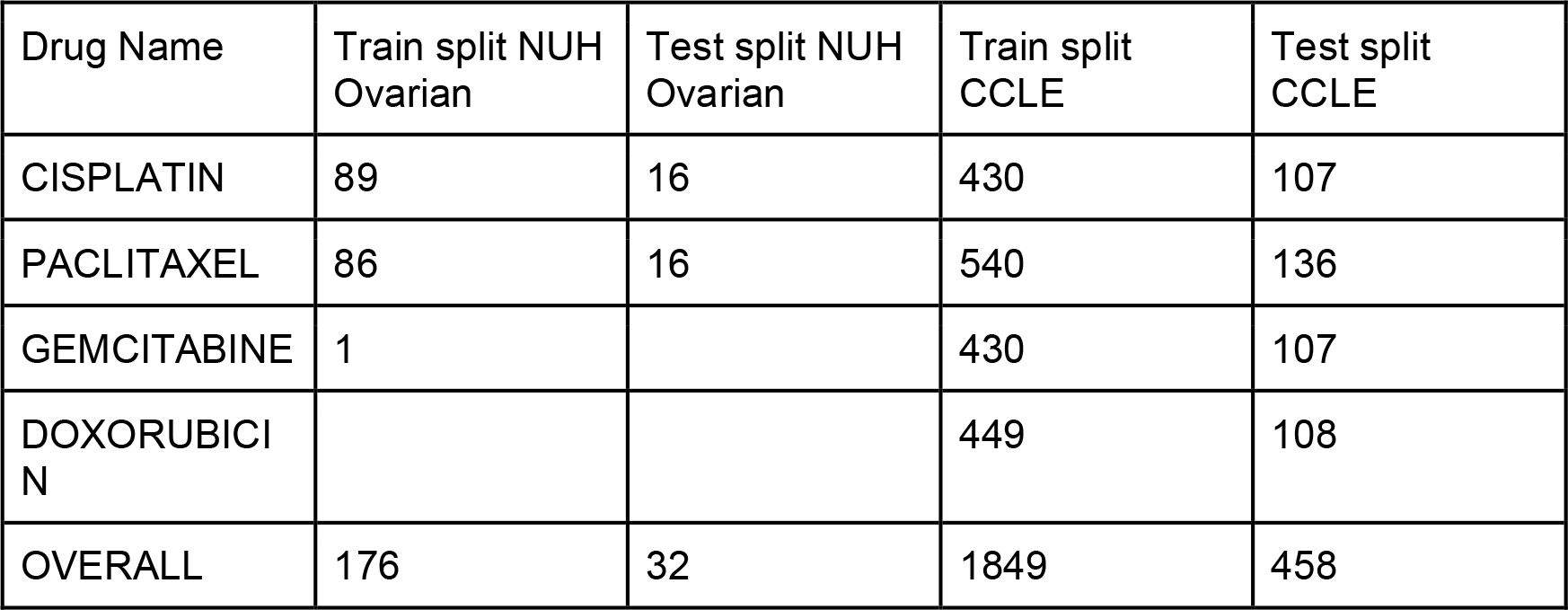
Number of (patient, drug) pairs in the train and test splits of CCLE-NUH Ovarian dataset split 2. Each cell indicates the number of patients/samples who were treated with the corresponding drug, which are further divided up across the train and test splits.

| Drug Name | Train split NUH Ovarian | Test split NUH Ovarian | Train split CCLE | Test split CCLE |
| --- | --- | --- | --- | --- |
| CISPLATIN | 89 | 16 | 430 | 107 |
| PACLITAXEL | 86 | 16 | 540 | 136 |
| GEMCITABINE | 1 |  | 430 | 107 |
| DOXORUBICIN |  |  | 449 | 108 |
| OVERALL | 176 | 32 | 1849 | 458 |

### Features

**Supplementary Table 16:** Summary of various input data types used across all the experiments in this paper. Mutations, copy number variations, gene expression and annotated mutation vectors are the key input data types. We further create different subsets based on the number of genes considered in each case. The resulting dimensions of the feature vectors are also described, in each case.

| Input Type | Feature Set | Encoding |
| --- | --- | --- |
| Mutations (F1 genes) vector | 324 genes sequenced in FoundationOne report | 324-dimensional binary vector with 1 bit per gene sequenced. A value of 1 indicates presence of mutation in a gene and 0 indicates its absence. |
| Mutations (All genes) vector | All 19536 genes sequenced | 19536-dimensional binary vector with 1 bit per gene sequenced. A value of 1 indicates presence of mutation in a gene and 0 indicates its absence. |
| Mutations (285 genes) vector | 285 genes sequenced in FoundationOne, Tempus xF+ and TruSight Oncology 500 reports | 285-dimensional binary vector with 1 bit per gene sequenced. A value of 1 indicates presence of mutation in a gene and 0 indicates its absence. |
| Gene expression (F1 genes) vector | 324 genes sequenced in FoundationOne report | 324-dimensional real-valued vector with 1 dimension per gene sequenced. No additional encoding done over raw data. |
| Copy number variation CNV (F1 genes) vector | 324 genes sequenced in FoundationOne report | 972-dimensional binary vector with 3 bits per gene sequenced. Raw data encoded with -1 indicating loss, +1 indicating amplification and 0 indicating no change. This was further one hot encoded to obtain 3 bits per gene. |
| Combined CNV and annotated mutation (F1 genes) vector | 324 genes sequenced in FoundationOne report | Obtained by concatenating VAE encoded representation for mutations (F1 genes) vector and VAE encoded representation for copy number variation CNV (F1 genes) vector. |
| Combined gene expression and annotated mutation (F1 genes) vector | 324 genes sequenced in FoundationOne report | 8100-dimensional vector obtained by concatenating annotated mutations (F1 genes) vector and gene expression (F1 genes) vector. |
| Variant annotated mutation (F1 genes) vector | 324 genes sequenced in FoundationOne report | 7776-dimensional vector obtained after variant annotation using Annovar, GPD and ClinVar, followed by an aggregation across all mutations in each gene. |

